# Designing a Clerkship Curriculum for Medical Students in Clinical and Medical Informatics in the Electronic Medical Record Era

**DOI:** 10.1101/2021.04.01.21253895

**Authors:** David Chartash, John T Finnell

## Abstract

**Introduction:** The nature of practice in clinical and medical informatics has changed since the last recommended incorporation of the field into undergraduate medical education in 1998. The Regenstrief Institute and Indiana University have been offering a clerkship in clinical and medical informatics since 2008, and post-graduate medical education since the early 1970s.

**Methods:** Drawing on this existing educational program and the expertise at the institute, we developed a clerkship elective to support medical students in learning about the subspecialty in practice, as well as the state of the art in research. Student evaluation of the clerkship and satisfaction with curricular design elements was measured.

**Results:** A modular inverted classroom curriculum was designed and implemented using learning management systems and instructional technology available at Indiana University. Students were satisfied with the clerkship, and explicitly understood how curricular design contributed to their understanding of material. The curriculum was found flexible for multiple levels of learners, particularly those with advanced mastery of technical skills.

**Discussion:** This curriculum, and its packaged content to be imported into common learning management systems, offers a state of the art introduction to clinical and medical informatics for medical students, which supports student success and preferences during their clerkship elective period.

**Educational Objectives:** By the end of this activity, learners will be able to:’

1. Articulate the scope of clinical and medical informatics within medicine
2. Describe a problem pertinent to their clinical interests, and how a solution can be supported by health information technology, as well as more generally be assessed using clinical outcome measures
3. Demonstrate a knowledge of terminologies and standards found in clinical medicine, as well how they integrate to define and describe disease
4. Demonstrate an understanding of clinical decision making and the evaluation of discrete choice problems

## Introduction

From the Medical School Objectives Project^1^ and recent curriculum renewal efforts by the American Medical Association (Accelerating Change in Medical Education Consortium), medical informatics has become an increasingly pertinent discipline of medicine in the undergraduate, graduate and post-graduate education of physicians. Medical informatics, however, has not been taught significantly in undergraduate medical education due to the lack of curricular space, increasing interest in topics that crowd out informatics (such as those which comprise the remainder of health systems science), and local failure of curricular reform. Rather, schools have provided elective opportunities in medical informatics for clerkship students looking to explore the discipline, particularly as it pertains to sub-specialty practice and board certification in clinical informatics. ^2^ In this vein, Indiana University School of Medicine and the Regenstrief Institute have been running a fourth/third year clerkship program since 2008, teaching fundamentals of medical informatics in a contained one-month experience covering four content areas: clinical decision support, coding and terminology, evidence-based medicine and statistics, and databases.^3^

Together, Indiana University School of Medicine and the Regenstrief Institute convened to create a clerkship and residency elective in medical informatics. The elective’s modules focused on research activates at the Institute and fundamental elements of medical informatics. The database module was facilitated through the use of a MySQL server within the Institute, and students were given access to a sandbox to play with an existing cohort of HIPAA complaint medical data. Due to infrastructure changes, as well as legal and policy mandates at the Institute, this become a less tenable option, and around 2011, students were instead directed to university-wide MySQL virtual instances and directed to perform tasks on data in their non-academic life (such as making a wedding registry). In addition to the changing landscape within the Institute, the overarching field of medical informatics had changed since the elective first began, with formal sub-speciality training and the emergent field of clinical informatics entering the lexicon of post-graduate medical education.^4–7^ Therefore, it was concluded that in preparation for the 2015-2016 academic year, the elective required a fundamental redesign to adapt to the changes in both medical and clinical informatics education.

The redesign of the curriculum was conducted during the 2014-2015 academic year. This coincided with the extension of the existing clinical informatics education at the Regenstrief institute to include an accredited clinical informatics fellowship between the Institute and medical school. Reviewing the competencies and curriculum suggested by *Silverman et al*^8^, as well as colleagues within the Institute, the core elements of the curriculum were redefined: fundamentals of medical informatics, decision science, terminologies and standards, final project in clinical decision support design, and the luminary paper series. In addition to these elements, two optional modules and one required pre-course module were developed. The former consists of a module on programming and databases, while the latter consists an overview of both Health Insurance Portability and Accountability Act (HIPAA) and key Federal legislation associated with reimbursement for health information technology (HIT). The decision to focus on three core modules, with a paper series delivered didactically was intended to provide a more flexible environment for students to complete residency interviews and USMLE exams. In addition, rather than holding students to a strict daily schedule, modules were rewritten to incorporate the inverted classroom pedagogical model. This was intended to support the engagement of clinical fellows with medical students during the limited didactic hours, as well as provide students with a clinically-focused approach to applying informatics and learning about the grand challenges from practicing clinical informaticians. The optional modules were provided for students with a technical background entering the course, as well as to support the use of the course as archived continuing education material during residency and beyond. The following provides a description of the curriculum, as well as the results from learner perception of the redesign compared to the previous curriculum as reported by *Finnell and Vreeman*^3^.

## Methods

The curriculum was initially designed for both medical students and house staff. The renewal efforts explicitly targets fourth year medical students, with a secondary target of third year, with the modularity focused on flexibility and asynchronous learning rather than enabling curricular growth over time. While previously students were expected to attend weekly classes and an extensive immersion in the Institute’s research activities, due to shifting priorities the latter was no longer a possibility. With the reduction in contact hours, a choice was made to shift the weekly sessions to provide contact with clinical informatics fellows alongside lecture material, and enable asynchronous learning. This contact with clinical informatics fellows allowed a direct experience with post-graduate education opportunities similar to rotations with services in which a clinical fellowship is required (such as critical care medicine). Furthermore, it supported the fellows’ development of educational competencies. Asynchronous learning was operationalized through an inverted classroom, and students were provided with the flexibility of connecting to lecture and tutorial sessions through telephone and video conference equipment. Didactic sessions each week were structured to consist of two one-hour tutorial sessions with clinical and informatics fellows, with a third hour of lecture by clinical faculty. The tutorial sessions offer opportunities for the fellows to lecture on clinical experiences of relevance to the weekly module in question, as well as answer questions in the context of the inverted classroom pertaining to said module. This supported students taking the United States Medical Licensing Examination (USMLE), as well as attending to residency interviews.

The curriculum was scaffolded around three central modules, with an optional fourth module. Each module was intended to be completed in a week’s time. Additional modules detailing gateway content, as well as longitudinal material filled out the curricular content. These longitudinal modules included summative assessments, optional modules to explore the depth and breadth of programming, and in the case of the luminary series, reading material for lectures. Although students were typically on a four-week block schedule, the compressed time frame allowed for a buffer of time to account for the aforementioned interviews, USMLE writing as well as final assessment completion. While the curricular content was designed to go into depth beyond the needs and wonts of a fourth year medical student, the material was designed to provide a basis for study during residency as well as for students entering with a more robust technical background (such as an undergraduate degree in biomedical engineering) an opportunity to stretch the limits of the material beyond the average medical student.

Figure 1 describes the sequence of modules and assessments, beginning with HIPAA Training and progressing in parallel through the sequence of modules, final project and luminary paper series.

**Figure 1.**
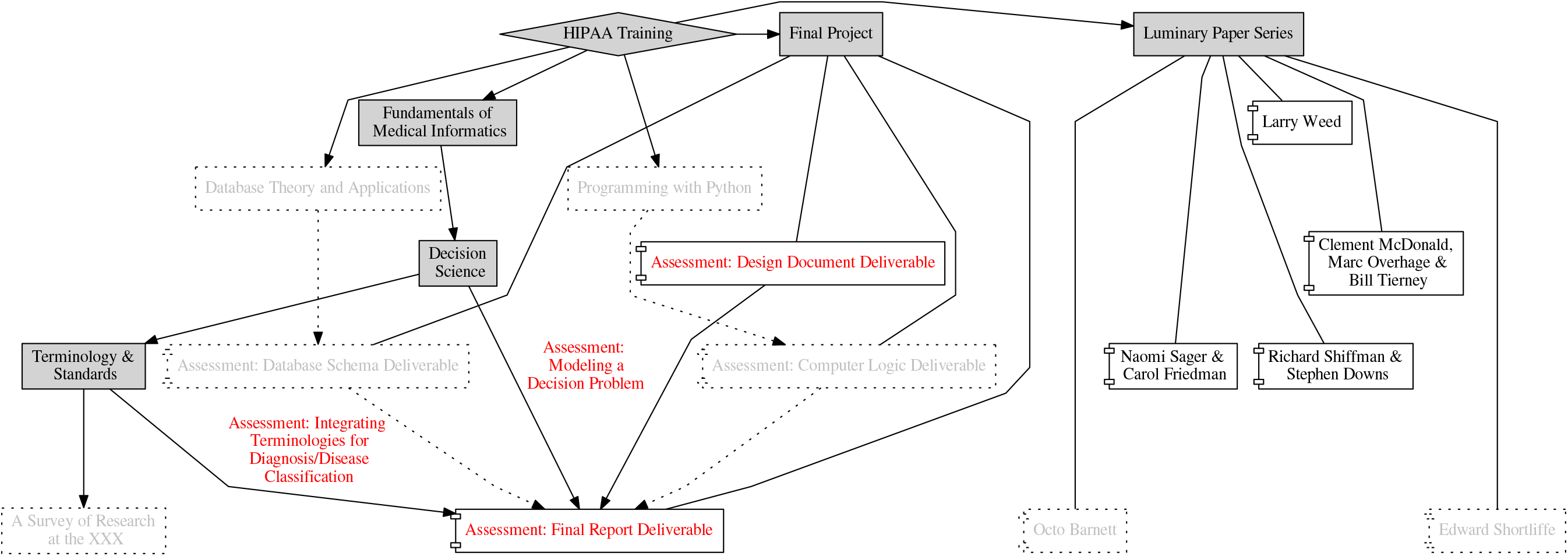
Diagram of module sequence. Optional modules are in grey silhouette, assessments in red, and required modules in dark grey.

**Table 1.** Table of module content, italicized content is optional.

| Module Name | Objectives | Weeks | Assessment |
| --- | --- | --- | --- |
| HIPAA Training | Educate through casework and university/institute material students regarding:<br>- The Health Information Technology for Economic and Clinical Health as a component of The American Recovery and Reinvestment Act<br>- The Health Insurance Portability and Accountability Act<br>- The Medicare Access and CHIP Reauthorization Act of the 114th Congress | 0 | - Quiz<br>- Attestation |
| Luminary Paper Series | This module introduces students to the grand challenges in medical informatics through the seminal papers of luminaries in the field:<br>- Lawrence L Weed: The Problem-based Medical Record and Electronic Medical Record<br>- Clement McDonald, J Marc Overhage and Bill Tierney: The Electronic Medical Record and Network, of Records -- Health Information Exchange<br>- Naomi Sager and Carol Friedman: Medical and Clinical Natural Language Processing<br>- Richard N Shiffman and Stephen M Downs: Clinical Guidelines and the Computer<br>*If the cohort of students is sufficiently advanced that they are capable of investigating artificial intelligence topics and database systems, two additional grand challenges and luminaries are kept in reserve:*<br>- Edward Shortliffe: *Artificial Intelligence and Expert Systems in Medicine*<br>- Octo Barnett: *The Origin of Medical Data and Timesharing Systems* | 0-4 | Lecture Participation |
| Final Project | Introduce students to the software engineering design process by walking them through designing a clinical | 0-4 | - Design Document deliverable<br>- *Computer Logic* |
|  | decision support system for their intended residency.<br>Specifically the deliverables include the tasks of:<br>1. Write a design document describing the clinical decision the student wishes to address, and the clinical problem it revolves around.<br>2. Design a database to describe the information sources capable of feeding into such a design solution.<br>3. Write in the Python programming language or pseudo-code the logic necessary to access and deploy the designed clinical decision support system.<br>4. Write a final report, detailing the solution as changed from design to final product, using the content now learned from the modules throughout the course. |  | *Deliverable*<br>- *Database Schema*<br>*Deliverable*<br>- Final Report<br>Deliverable |
| Programming with Python & Database Theory and Applications | Guide students through externally developed content in both database design and Python programming, both at a first year undergraduate level. | 0-4 | Optional deliverables per material |
| Fundamentals of Medical Informatics | Educate students regarding:<br>- The extension of the biopsychosocial model of disease into the cybernetic and information space and its subsequent evolution into the infomedical model<br>- Perspectives on the theory of medicine that influence clinical epidemiology and practice: clinical semiotics and Bayesian models of clinical reasoning<br>- The origins of medical informatics in the United States and its shaping by pioneers at the Regenstrief Institute as well as other members of the American College of Medical Informatics (ACMI)<br>- Clinical informatics and the intersection of care and computers, alongside the role of translational science and research: graduate medical education in informatics. | 1 | Tutorial Participation |
| Decision Science |  | 2 | Modeling a Decision |
|  | - Introduce the notion of the discrete choice experiment<br>- Describe the Bayesian cognitive model and how it integrates into probabilistic decision-modeling<br>- Provide a guided tour through decision science principles such as influence diagrams, decision trees, cost-effectiveness, cost-utility and cost-benefit analysis |  | Problem |
| Terminology and Standards | Introduce students to the use of terminologies and standards they will observe in clinical practice, and which are useful for the design of clinical decision support:<br>- Systematized Nomenclature of Human and Veterinary Medicine - Clinical Terms (SNOMED-CT)<br>- International Classification of Disease<br>- American Clinical Modification, Versions 9 and 10<br>- RxNorm, the US-centric normalized system for drug naming and semantic linkages between drug and pharmacy terminologies and knowledge.<br>- Health Level 7 International Messaging Version 2.8, Fast Healthcare Interoperability Resources Specification and Arden Syntax<br>- Logical Observation Identifiers Names and Codes (LOINC) | 3 | Integrating Terminologies for Diagnosis/Disease Classification |
| A Survey of Research at the Regenstrief Institute | This module is necessarily tailored to engage students in research activities within the department, center or institute offering the elective. At the Regenstrief Institute, this details research areas, faculty members, major projects, and examples of clinical informatics research that the students have seen during clerkship. | 4 | None |

The first module encountered by clerkship students is the HIPAA Training module. Students were expected to complete this module asynchronously prior to matriculation. The primary content was reading and legal casework concerning each of the three statutes pertinent to understanding the deployment of medical informatics and the use of data within the clinical environment for purposes other than clinical care: The Health Information Technology for Economic and Clinical Health as a component of The American Recovery and Reinvestment Act, The Health Insurance Portability and Accountability Act, and The Medicare Access and CHIP Reauthorization Act of the 114th Congress. Two assessments were used to provide a gateway between the HIPAA module and the remainder of the content. The first of these was modules from the Collaborative Institutional Training Initiative at the University of Miami administrated through the Indiana University Office of Research Administration: biomedical and social science researcher, responsible conduct of research and good clinical practice. The second was a quiz designed for the Regenstrief Institute as a covered entity. Successful completion of these two assessments with greater than an 80% score was considered a passing grade. Once this module’s assessments were completed, the subsequent modules were unlocked for the student to complete. The modules were discussed in tutorial and lectures in the sequence suggested by Figure 1.

The second module therefore, is entitled Fundamentals of Medical Informatics. This module contains readings detailing the discipline of medical informatics as it is currently practiced and defined by the community, as well as relates to existing knowledge within the undergraduate medical curriculum. Specifically, it covers the extension of the biopsychosocial model of disease into the cybernetic and information space and its subsequent evolution into the infomedical model; perspectives on the theory of medicine that influence clinical epidemiology and practice (specifically clinical semiotics and the Bayesian models of clinical reasoning); the origins of medical informatics in the United States and its shaping by pioneers at the Regenstrief Institute as well as other members of the American College of Medical Informatics (ACMI); and the creation of the sub-speciality of clinical informatics and the intersection of care and computers, alongside the role of translational science and research in graduate medical education. This module is not assessed through a quiz, but rather through the interaction between fellows and student during tutorial sessions. The intention is to ground students in theory and provide them with a reflective tie to prior curricular content that they have integrated by their fourth year into their armamentarium and demonstrated mastery over through the first step of the United States Medical Licensing Examination (USMLE).

The third module is an overview of Decision Science, and its application in medicine. First, this module re-introduces students to the fundamentals of both probability and Bayesian statistics that have been covered by prior curricular items, and expands these concepts to include information theory. By working through the primer series on medical decision analysis in the 17th volume Medical Decision Making (1997), students are introduced to the concepts of discrete choice, modeling through trees and the use of probabilities and utilities to evaluate medical decisions. This content is supplemented by a brief description adapted from clinical informatics board study materials^9^ along with seminal texts in health economics. To assess their knowledge of the material, students are asked to model a clinical decision (and are suggested to select one that aligns with their final project) of interest to their clinical practice intent. Using this model, students are asked to roughly quantify the effect of different elements of the decision, as would be done in a formal decision analysis. Given that data sources and the lack of quantification is a major barrier to performing decision analysis (irrespective of the topic), students are expected to run into this barrier and learn about the lack of evidence to formally evaluate what they often perceive as solved or known problems.

The fourth module is an overview of Terminologies and Standards present in medicine, focusing on diagnosis and the electronic medical record. This module introduces students to the Systematized Nomenclature of Human and Veterinary Medicine - Clinical Terms (SNOMED-CT); International Classification of Disease - American Clinical Modification, Versions 9 and 10; RxNorm, the US-centric normalized system for drug naming and semantic linkages between drug and pharmacy terminologies and knowledge; Health Level 7 International Messaging Version 2.8, Fast Healthcare Interoperability Resources Specification and Arden Syntax; and Logical Observation Identifiers Names and Codes (LOINC). In addition, students are introduced to the Unified Medical Language System Terminology Services, through which the connectivity and semantics of the various terminologies and standards are presented. Students are assessed by asking them to expand on their knowledge of these standards and the terminology service by selecting a clinical concept pertinent to their intended clinical practice (again, it is suggested that they align this concept with their final project) and explain how the differences between each of the terminologies and standards come together to provide a more complete picture of the disease. In addition, students are asked to hypothesize how this more complete picture (and the individual terminologies and standards) influence clinical practice, either directly or indirectly.

In addition to these four modules that are core components of the clerkship, a module introducing students to research at the Regenstrief Institute (or in duplication, the department, center or institute offering the clerkship) was developed. This was simply a description of major efforts, projects and teams at the Institute, conveyed through a listing of projects and papers as well as recorded grand rounds and other presentations. This provided a basis for students who were intending to stay at Indiana for residency, or were completing additional scholarly components during a fifth year (such as a masters in public health or masters in clinical research) to become familiar with the Institute’s research for future collaboration.

Beyond these five modules that exist as weekly or discrete content, three modules provide longitudinal material throughout the clerkship. The first of these modules contains the didactic content that is the basis of the luminary series lectures each week. The grand challenges of informatics: natural language processing, Electronic Medical Record (EMR) design and health information exchange, clinical guidelines and decision support, and artificial intelligence are discussed through the lens of the seminal authors in the field. For natural language processing, a survey of the work of Naomi Sager and Carol Friedman is provided. For EMR design and health information exchange, separately, the work of Clement McDonald, J Marc Overhage, Bill Tierney, database work of Octo Barnett and problem-oriented medical record design work of Lawrence L Weed is discussed. For clinical guidelines and decision support the work of both Stephen M Downs and Richard N Shiffman is discussed. For artificial intelligence, the papers of Edward Shortliffe are discussed. These papers allow students to compare the current state of the art as related in lectures and popular media, to the seminal literature, and identify how much progress has been made over the years to meet the grand challenge in question. As there are more sets of luminaries than there are weeks in the clerkship, typically Edward Shortliffe and Octo Barnett’s papers are provided as optional reading. While both luminaries are important to informatics, they are less important to the student entering residency, the technical complexity of artificial intelligence has prevented it from being used at the bedside, and Octo Barnett’s development of MUMPS is pertinent in the context of interacting deeply with the EMR, a task not typically undertaken in residency.

The second module is an optional module collecting content on programming with the Python language, as well as the basics of database design and theory. This is provided as optional content, given the level of engagement of students to the clerkship. Given the light workload compared to clinical electives, it often fosters weak engagement (as demonstrated by 3), and therefore the programming and database content was left as optional depth. Additionally, this provides students with prior knowledge a platform with which to practice their skill-set, which inevitably has been dulled by the past three years of medical school. Programming content is adapted from the work of 10. The database content is adapted from 11 as implemented on Stanford’s edX platform.

The final module is the summative activity for the clerkship. It begins with a discussion of the notion of engineering design, and how a design document is structured. Students are expected to complete the following deliverables, with the database schema and computer logic optional unless the student has prior experience or has completed the associated module:

1. *Design Document:* Write a design document describing the clinical decision the student wishes to address, and the clinical problem it revolves around.
2. *Database Schema:* Design a database to describe the information sources capable of feeding into such a designed solution. It is suggested that students use the following as a guide:
  1. Given the OpenMRS Data Model, describe what data is required by the application.
  2. Use Unified Modeling Language (UML) to detail the functional needs of the problem, given objects from the OpenMRS data model.
3. *Computer Logic:* Write in Python (or pseudo-code) the code necessary to access the described database/data model. This code should provide the clinical logic at point of care that would accompany the system designed in the hospital/clinical environment that has been defined.
4. *Final Report:* Write a final report, detailing as much of the solution as possible. Specifically include:
  1. Detail the design of the decision support system, as well as the rationale for each component given the clinical circumstances
  2. Describe the clinical circumstances of the problem, and ponder hypotheses that this solution would be capable of influencing or testing. This report should integrate the knowledge gained throughout this course, specifically that from other modules to supplement the design document in this final report.

The final project provides the clerkship instructors an assessment of meta-cognitive capacity, and the depth of the students’ knowledge gained during the course. The assessment is designed such that if a student has sufficient expertise a functioning decision support system can be designed and developed during the month. While this did not take place in the years since the curriculum renewal, students have developed functional decision support systems capable of accepting user input and acting upon them extraneous from the EMR. The deliverables are sequentially tied to the first through fourth week, as this provides adequate time for learning of the content if they pursue all of the optional modules. In addition, the design document is necessarily completed prior to the final report.

Following the completion of the clerkship rotation, students evaluated the clerkship using the forms standard to the medical school. These evaluations were compared against the previous evaluations to identify areas of strength, improvement, student preferences, and satisfaction with curriculum and renewal efforts.

## Results

The renewed clerkship curriculum was finalized for the 2014-2015 academic year, and has been offered on a monthly basis. Enrollment between 2014 and 2018 has averaged 30 students per year (an average of 8.24% of the medical student class), with enrollment decreasing over time. Of these students, an average of 22 students filled out the evaluations yearly. These numbers are compared to 20 students having taken the elective from 2008-2010. Students’ recognition of the value of learning about medical informatics in an elective experience is suggested as the rationale for an increased attendance. The decrease from the 2015 academic year is most likely due to an online elective in health information technology offered at the Terre Haute campus by an adjunct lecturer.

The mean overall rating of the elective was 3.34, on a four point likert scale. This is an incremental improvement over the previous evaluation average of 4 on a five point scale (poor, fair, good, very good, excellent). Comparison of qualitative comments regarding the clerkship suggested that it provided the same level of exposure and novel knowledge. The increase in depth of material between the prior course and the renewed curriculum was initially taken askance, however, over time, communication of the syllabus and required effort from the students was improved. This resulted in the independent comment of students that the course was intended to build upon its own material through summative assessments and sequenced modular content. The increase in rigor compared with prior coursework (such as it related to the inverted classroom and independent reading) made this elective difficult for medical students who engaged with the bare minimum of the material.

In general, however, the renewed curriculum was praised for its flexibility and modularity introducing content to clinically-focused students. Students found direct interaction with clinical informatics fellows valuable in developing an understanding of medical informatics at point-of-care. Course faculty and fellows were often approached after the clerkship had concluded to engage for future research and education during residency.

## Discussion

Our curriculum renewal was successful in several ways. First, we incrementally increased student satisfaction with the elective while increasing the rigor of the curriculum. Second, the flexibility and modularity of the curriculum was optimized, a lauded improvement from the students’ perspective. Third, in concert with the clinical informatics fellowship, the renewal focused the clinical applicability of the elective through summative assessment. This resulted in students understanding that the material was introductory yet constructive, building on their clinical knowledge and the remainder of their medical school curriculum to support future practice.

While similar curricula emphasize research engagement or clinical shadowing^2^, we found that a mixed focus that emphasized further post-graduate practice in clinical emphasis was the optimal theme for curriculum renewal. Furthermore, the use of an inverted classroom supports students’ activities in fourth year. While explicit value of the luminary series reflecting grand challenges, as compared to module content providing fundamentals was not realized, in general the connectivity of the modular structure of the curriculum was extremely successful in guiding students to understand how the material enhances prior clinical knowledge for future practice. Furthermore, while similar curricula have been developed and compared among the clinical and medical informatics education community, there is a consistent lack of standards in this portion of undergraduate medical education. We propose that our curriculum offers a starting point for discussions, alongside the shareable content in Appendix A.

Limitations in the curriculum included political and logistical challenges. First, given that the medical school allows competing electives as long as education requirements are met, a competing online only elective in Terre Haute siphoned student enrollment. Students make decisions based on several factors when selecting electives, and an a solely online elective out performs our curriculum logistically. Furthermore, the enhancement of rigor was seen by students as a disadvantage for an elective that would not immediately advance their clinical skills. Decreasing enrollment is a concern for the elective in the future, however, efforts to combat such attrition will include advertisement with interest groups within the school. Logistically, beyond the Institute’s administrative challenges, communication with medical students from clinician educators while the students were not engaged with their service lines resulted in difficulty implementing the inverted classroom.

## Data Availability

The data is available from the authors at the reader's request.

## Appendices

A. <u>Canvas_Modules.zip</u>

