## Supplemental File and Appendix for "Designing a Clerkship Curriculum for Medical Students in Clinical and Medical Informatics in the Electronic Medical Record Era": Designing_MedRxiv_ESR_2018_CanvasSummary.docx

### Appendix A: Course Content

The course begins with the syllabus (Syllabus.docx), which describes the overall structure of the course material detailed in the ESR, as well as the expectations of students. Prior to the course, students are asked to complete a pre-course questionnaire. This questionnaire is provided within the Canvas LMS, and therefore is shown as an HTML form (Pre-Course_Questionnaire.html), we have also provided a PDF capture of the static form (Pre-Course_Questionnaire.pdf).

The overarching content of the programming and database modules (Programming (with Python) and Databases - Pages Concatenated.docx) detail the location of material and expected assignment completion (questions for chapters or modules within the edX instance). The final project module details the expected deliverables which are aligned with the syllabus (Final Project - Pages Concatenated.docx). The luminary paper series module (Luminary Paper Series - Pages Concatenated.docx) details the week-long blocks, to be selected by instructors that fit into the course schedule. Each luminary (or luminaries) comprises a week-long block, and has a list of reading prior to the discussion during the week. For most reading material, those articles which are starred are the seminal articles for the week, however, in some cases students are expected to read all of the material (in particular, Larry Weed).

Starting the course, students are expected to complete the HIPAA training, covered in the module content (HIPAA Training - Pages Concatenated.docx) and complete the quiz, similarly delivered via Canvas and provided as an HTML form (HIPAA_Quiz.html) and a PDF capture of the static form (HIPAA_Quiz.pdf). The sequenced modules detail the reading and summary material expected of the student to engage with instructors, as well as assignments for the module (see Fundamentals of Medical Informatics - Pages Concatenated.docx, Decision Science - Pages Concatenated.docx, Terminologies and Standards - Pages Concatenated.docx). The survey of research module (A Survey of Regenstrief's Current Research - Pages Concatenated.docx) is a breakdown of the research aligned with course content around the university at time of reporting. It is provided as an optional module for students intending to stay at Indiana for residency, as well as those taking it and interested in future research. We have provided this module as a template for how other institutions may present their areas of expertise and points of engagement for medical students.

In running the course, we would recommend uploading the readings available digitally within the university's library to the LMS. Some material (especially that which is out of copyright) may be accessible through the HathiTrust Digital Library.
