## Supplemental File and Appendix for "Designing a Clerkship Curriculum for Medical Students in Clinical and Medical Informatics in the Electronic Medical Record Era": HIPAA_Quiz.html

Quiz: Regenstrief HIPAA Quiz


### You need to have JavaScript enabled in order to access this site.


Close

##### Loading...

### Regenstrief HIPAA Quiz

This is a preview of the published version of the quiz

Started:
Jan 30 at 9:18pm

#### Quiz Instructions

You must complete this quiz with a mark of 80% or greater to continue with the remainder of this course.
  
You may take this quiz as many times as you desire, until this score is met. You may only view your responses once after each attempt.


2019-01-30T21:18:24-05:00

2019-01-30T21:18:24-05:00

2060-06-29T22:59:00-05:00


1306892435

*Move To...

This element is a more accessible alternative to drag & drop reordering. Press Enter or Space to move this question.*

Flag this Question

Question 1


1
pts

*Edit this Question* 

 *Delete this Question*

0

multiple\_choice\_question

Which of the following is appropriate for Social Media?

Which of the following is appropriate for Social Media?

|  |  |
| --- | --- |
|  | None of the above |

|  |  |
| --- | --- |
|  | Private Conversations pertaining to Intellectual Property, Confidential Information, or other proprietary information |

|  |  |
| --- | --- |
|  | New ideas that include Intellectual Property |

|  |  |
| --- | --- |
|  | All of the above |

|  |  |
| --- | --- |
|  | Confidential Information |

|  |  |
| --- | --- |
|  | PHI |

Next

2019-01-30T21:18:24-05:00

2019-01-30T21:24:44-05:00

2060-06-29T22:59:00-05:00


1306892056

*Move To...

This element is a more accessible alternative to drag & drop reordering. Press Enter or Space to move this question.*

Flag this Question

Question 2


1
pts

*Edit this Question* 

 *Delete this Question*

0

true\_false\_question

If I find a USB, I should plug it into my computer to find the owner.

If I find a USB, I should plug it into my computer to find the owner.

|  |  |
| --- | --- |
|  | True |

|  |  |
| --- | --- |
|  | False |

Previous


Next

2019-01-30T21:18:24-05:00

2019-01-30T21:24:49-05:00

2060-06-29T22:59:00-05:00


1306892051

*Move To...

This element is a more accessible alternative to drag & drop reordering. Press Enter or Space to move this question.*

Flag this Question

Question 3


1
pts

*Edit this Question* 

 *Delete this Question*

0

true\_false\_question

I can give my password out as long as it is to someone in IT.

I can give my password out as long as it is to someone in IT.

|  |  |
| --- | --- |
|  | True |

|  |  |
| --- | --- |
|  | False |

Previous


Next

2019-01-30T21:18:24-05:00

2019-01-30T21:25:02-05:00

2060-06-29T22:59:00-05:00


1306892038

*Move To...

This element is a more accessible alternative to drag & drop reordering. Press Enter or Space to move this question.*

Flag this Question

Question 4


1
pts

*Edit this Question* 

 *Delete this Question*

0

multiple\_choice\_question

Social engineering can be done via:

Social engineering can be done via:

|  |  |
| --- | --- |
|  | Hyperlinks |

|  |  |
| --- | --- |
|  | Text |

|  |  |
| --- | --- |
|  | Dropped media (CDs, USBs, etc.) |

|  |  |
| --- | --- |
|  | Email |

|  |  |
| --- | --- |
|  | In person |

|  |  |
| --- | --- |
|  | All of the above |

|  |  |
| --- | --- |
|  | None of the above |

Previous


Next

2019-01-30T21:18:24-05:00

2019-01-30T21:25:07-05:00

2060-06-29T22:59:00-05:00


1306892032

*Move To...

This element is a more accessible alternative to drag & drop reordering. Press Enter or Space to move this question.*

Flag this Question

Question 5


1
pts

*Edit this Question* 

 *Delete this Question*

0

true\_false\_question

Connecting smartphones and tablets to Regenstrief email requires accepting the Regenstrief security policies including a six digit pin.

Connecting smartphones and tablets to Regenstrief email requires accepting the Regenstrief security policies including a six digit pin.

|  |  |
| --- | --- |
|  | True |

|  |  |
| --- | --- |
|  | False |

Previous


Next

2019-01-30T21:18:24-05:00

2019-01-30T21:25:14-05:00

2060-06-29T22:59:00-05:00


1306892026

*Move To...

This element is a more accessible alternative to drag & drop reordering. Press Enter or Space to move this question.*

Flag this Question

Question 6


1
pts

*Edit this Question* 

 *Delete this Question*

0

true\_false\_question

PHI may not be stored on laptops, tablets, cell phones, or other mobile devices, even if the laptop, tablet, cell phone, or other mobile device is encrypted.

PHI may not be stored on laptops, tablets, cell phones, or other mobile devices, even if the laptop, tablet, cell phone, or other mobile device is encrypted.

|  |  |
| --- | --- |
|  | True |

|  |  |
| --- | --- |
|  | False |

Previous


Next

2019-01-30T21:18:24-05:00

2019-01-30T21:25:18-05:00

2060-06-29T22:59:00-05:00


1306892022

*Move To...

This element is a more accessible alternative to drag & drop reordering. Press Enter or Space to move this question.*

Flag this Question

Question 7


1
pts

*Edit this Question* 

 *Delete this Question*

0

true\_false\_question

Regenstrief is not subject to audit for HIPAA Compliance because it is a Business Associate and not a Covered Entity.

Regenstrief is not subject to audit for HIPAA Compliance because it is a Business Associate and not a Covered Entity.

|  |  |
| --- | --- |
|  | True |

|  |  |
| --- | --- |
|  | False |

Previous


Next

2019-01-30T21:18:24-05:00

2019-01-30T21:25:22-05:00

2060-06-29T22:59:00-05:00


1306892018

*Move To...

This element is a more accessible alternative to drag & drop reordering. Press Enter or Space to move this question.*

Flag this Question

Question 8


1
pts

*Edit this Question* 

 *Delete this Question*

0

true\_false\_question

Email should NOT be forwarded to personal email accounts.

Email should NOT be forwarded to personal email accounts.

|  |  |
| --- | --- |
|  | True |

|  |  |
| --- | --- |
|  | False |

Previous


Next

2019-01-30T21:18:24-05:00

2019-01-30T21:25:26-05:00

2060-06-29T22:59:00-05:00


1306892014

*Move To...

This element is a more accessible alternative to drag & drop reordering. Press Enter or Space to move this question.*

Flag this Question

Question 9


1
pts

*Edit this Question* 

 *Delete this Question*

0

true\_false\_question

All IU programs and services offered through UITS are appropriate for communicating PHI.

All IU programs and services offered through UITS are appropriate for communicating PHI.

|  |  |
| --- | --- |
|  | True |

|  |  |
| --- | --- |
|  | False |

Previous


Next

2019-01-30T21:18:24-05:00

2019-01-30T21:25:30-05:00

2060-06-29T22:59:00-05:00


1306892010

*Move To...

This element is a more accessible alternative to drag & drop reordering. Press Enter or Space to move this question.*

Flag this Question

Question 10


1
pts

*Edit this Question* 

 *Delete this Question*

0

true\_false\_question

When accessing PHI, I should only access the minimum that is necessary to do my job. This means not accessing information I find interesting or about friends, family members, coworkers, etc.

When accessing PHI, I should only access the minimum that is necessary to do my job. This means not accessing information I find interesting or about friends, family members, coworkers, etc.

|  |  |
| --- | --- |
|  | True |

|  |  |
| --- | --- |
|  | False |

Previous


Next

2019-01-30T21:18:24-05:00

2019-01-30T21:25:34-05:00

2060-06-29T22:59:00-05:00


1306892005

*Move To...

This element is a more accessible alternative to drag & drop reordering. Press Enter or Space to move this question.*

Flag this Question

Question 11


1
pts

*Edit this Question* 

 *Delete this Question*

0

multiple\_choice\_question

A good definition of Social Engineering can include:

A good definition of Social Engineering can include:

|  |  |
| --- | --- |
|  | "tricking people to get them to do what you want them to do" |

|  |  |
| --- | --- |
|  | All of the above |

|  |  |
| --- | --- |
|  | "any act that influences a person to take an action that may or may not be in their best interest" |

|  |  |
| --- | --- |
|  | "the manipulation of social position and function of individuals in order to manage change in society" |

Previous


Next

2019-01-30T21:18:24-05:00

2019-01-30T21:25:38-05:00

2060-06-29T22:59:00-05:00


1306892001

*Move To...

This element is a more accessible alternative to drag & drop reordering. Press Enter or Space to move this question.*

Flag this Question

Question 12


1
pts

*Edit this Question* 

 *Delete this Question*

0

true\_false\_question

All paper should be disposed in PHI bins.

All paper should be disposed in PHI bins.

|  |  |
| --- | --- |
|  | True |

|  |  |
| --- | --- |
|  | False |

Previous


Next

2019-01-30T21:18:24-05:00

2019-01-30T21:25:43-05:00

2060-06-29T22:59:00-05:00


1306891996

*Move To...

This element is a more accessible alternative to drag & drop reordering. Press Enter or Space to move this question.*

Flag this Question

Question 13


1
pts

*Edit this Question* 

 *Delete this Question*

0

true\_false\_question

PHI can be emailed if it is to someone within Regenstrief, Eskenazi, or IU Health, and the subject line includes the words "secure message" or "confidential."

PHI can be emailed if it is to someone within Regenstrief, Eskenazi, or IU Health, and the subject line includes the words "secure message" or "confidential."

|  |  |
| --- | --- |
|  | True |

|  |  |
| --- | --- |
|  | False |

Previous


Next

2019-01-30T21:18:24-05:00

2019-01-30T21:25:49-05:00

2060-06-29T22:59:00-05:00


1306891990

*Move To...

This element is a more accessible alternative to drag & drop reordering. Press Enter or Space to move this question.*

Flag this Question

Question 14


1
pts

*Edit this Question* 

 *Delete this Question*

0

multiple\_choice\_question

In order to assist with HIPAA compliance, I can:

In order to assist with HIPAA compliance, I can:

|  |  |
| --- | --- |
|  | All of the above |

|  |  |
| --- | --- |
|  | Get to know my coworkers |

|  |  |
| --- | --- |
|  | Ensure the confidentiality statement is on my email signature |

|  |  |
| --- | --- |
|  | Be aware of my surroundings |

|  |  |
| --- | --- |
|  | Always lock my computer when I'm not using it |

Previous


Next

2019-01-30T21:18:24-05:00

2019-01-30T21:25:54-05:00

2060-06-29T22:59:00-05:00


1306891986

*Move To...

This element is a more accessible alternative to drag & drop reordering. Press Enter or Space to move this question.*

Flag this Question

Question 15


1
pts

*Edit this Question* 

 *Delete this Question*

0

true\_false\_question

Human error is the biggest concern for employers when thinking about a potential breach.

Human error is the biggest concern for employers when thinking about a potential breach.

|  |  |
| --- | --- |
|  | True |

|  |  |
| --- | --- |
|  | False |

Previous


Next

2019-01-30T21:18:24-05:00

2019-01-30T21:25:59-05:00

2060-06-29T22:59:00-05:00


1306891981

*Move To...

This element is a more accessible alternative to drag & drop reordering. Press Enter or Space to move this question.*

Flag this Question

Question 16


1
pts

*Edit this Question* 

 *Delete this Question*

0

true\_false\_question

Even a light bulb can be hacked.

Even a light bulb can be hacked.

|  |  |
| --- | --- |
|  | True |

|  |  |
| --- | --- |
|  | False |

Previous


Next

2019-01-30T21:18:24-05:00

2019-01-30T21:26:04-05:00

2060-06-29T22:59:00-05:00


1306891975

*Move To...

This element is a more accessible alternative to drag & drop reordering. Press Enter or Space to move this question.*

Flag this Question

Question 17


1
pts

*Edit this Question* 

 *Delete this Question*

0

true\_false\_question

HIPAA is the only limitation on how we use or share protected health information.

HIPAA is the only limitation on how we use or share protected health information.

|  |  |
| --- | --- |
|  | True |

|  |  |
| --- | --- |
|  | False |

Previous


Next

2019-01-30T21:18:24-05:00

2019-01-30T21:26:08-05:00

2060-06-29T22:59:00-05:00


1306891971

*Move To...

This element is a more accessible alternative to drag & drop reordering. Press Enter or Space to move this question.*

Flag this Question

Question 18


1
pts

*Edit this Question* 

 *Delete this Question*

0

true\_false\_question

If a laptop is stolen, destroyed by a sledge hammer, and thrown in a lake, it does not need to be reported to Legal and Compliance.

If a laptop is stolen, destroyed by a sledge hammer, and thrown in a lake, it does not need to be reported to Legal and Compliance.

|  |  |
| --- | --- |
|  | True |

|  |  |
| --- | --- |
|  | False |

Previous


Next

2019-01-30T21:18:24-05:00

2019-01-30T21:26:14-05:00

2060-06-29T22:59:00-05:00


1306891965

*Move To...

This element is a more accessible alternative to drag & drop reordering. Press Enter or Space to move this question.*

Flag this Question

Question 19


0
pts

*Edit this Question* 

 *Delete this Question*

0

essay\_question

My electronic signature below acknowledges that I have completed the Regenstrief Institute HIPAA compliance training. I agree to maintain the policies and standards of HIPAA compliance and fully participate with the Regenstrief Institute's compliance program.

My electronic signature below acknowledges that I have completed the Regenstrief Institute HIPAA compliance training. I agree to maintain the policies and standards of HIPAA compliance and fully participate with the Regenstrief Institute's compliance program.

HTML Editor

Rich Content Editor

Previous

Time's Up! Submitting results in:

Ok, fine

Not saved

Submit Quiz


**You have been logged out of canvas.** 
  
To continue please log in

88c612c7-0e76-4b83-8777-ac5c6990da79
