## Supplemental File and Appendix for "Designing a Clerkship Curriculum for Medical Students in Clinical and Medical Informatics in the Electronic Medical Record Era": HIPAA_Quiz.pdf

### Regenstrief HIPAA Quiz

#### Quiz Instructions

You must complete this quiz with a mark of 80% or greater to continue with the remainder of this course. You may take this quiz as many times as you desire, until this score is met. You may only view your responses once after each attempt.

---

**Question 1** **1 pts**

Which of the following is appropriate for Social Media?

- None of the above
- Private Conversations pertaining to Intellectual Property, Confidential Information, or other proprietary information
- New ideas that include Intellectual Property
- All of the above
- Confidential Information
- PHI

Next ▸

**Question 2** **1 pts**

If I find a USB, I should plug it into my computer to find the owner.

- True
- False

◂ Previous

Next ▸

**Question 3** **1 pts**

I can give my password out as long as it is to someone in IT.

- True
- False

◂ Previous

Next ▸

**Question 4** **1 pts**

Social engineering can be done via:

- Hyperlinks
- Text
- Dropped media (CDs, USBs, etc.)
- Email
- In person
- All of the above

- None of the above

◂ Previous　　Next ▸

#### Question 5 — 1 pts

Connecting smartphones and tablets to Regenstrief email requires accepting the Regenstrief security policies including a six digit pin.

- True
- False

◂ Previous　　Next ▸

#### Question 6 — 1 pts

PHI may not be stored on laptops, tablets, cell phones, or other mobile devices, even if the laptop, tablet, cell phone, or other mobile device is encrypted.

- True
- False

◂ Previous　　Next ▸

#### Question 7 — 1 pts

Regenstrief is not subject to audit for HIPAA Compliance because it is a Business Associate and not a Covered Entity.

- True
- False

◂ Previous | Next ▸

#### Question 8 — 1 pts

Email should NOT be forwarded to personal email accounts.

- True
- False

◂ Previous | Next ▸

#### Question 9 — 1 pts

All IU programs and services offered through UITS are appropriate for communicating PHI.

- True
- False

◂ Previous

Next ▸

#### Question 10

**1 pts**

When accessing PHI, I should only access the minimum that is necessary to do my job. This means not accessing information I find interesting or about friends, family members, coworkers, etc.

- True
- False

◂ Previous

Next ▸

#### Question 11

**1 pts**

A good definition of Social Engineering can include:

- "tricking people to get them to do what you want them to do"
- All of the above
- "any act that influences a person to take an action that may or may not be in their best interest"
- "the manipulation of social position and function of individuals in order to manage change in society"

◂ Previous

Next ▸

#### Question 12

**1 pts**

All paper should be disposed in PHI bins.

- True
- False

◂ Previous

Next ▸

#### Question 13

**1 pts**

PHI can be emailed if it is to someone within Regenstrief, Eskenazi, or IU Health, and the subject line includes the words "secure message" or "confidential."

- True
- False

◂ Previous

Next ▸

#### Question 14 — 1 pts

In order to assist with HIPAA compliance, I can:

- All of the above
- Get to know my coworkers
- Ensure the confidentiality statement is on my email signature
- Be aware of my surroundings
- Always lock my computer when I'm not using it

◂ Previous    Next ▸

#### Question 15 — 1 pts

Human error is the biggest concern for employers when thinking about a potential breach.

- True
- False

◂ Previous    Next ▸

#### Question 16 — 1 pts

Even a light bulb can be hacked.

- True

False

◂ Previous

Next ▸

#### Question 17

**1 pts**

HIPAA is the only limitation on how we use or share protected health information.

- True
- False

◂ Previous

Next ▸

#### Question 18

**1 pts**

If a laptop is stolen, destroyed by a sledge hammer, and thrown in a lake, it does not need to be reported to Legal and Compliance.

- True
- False

◂ Previous

Next ▸

#### Question 19

0 pts

My electronic signature below acknowledges that I have completed the Regenstrief Institute HIPAA compliance training. I agree to maintain the policies and standards of HIPAA compliance and fully participate with the Regenstrief Institute's compliance program.

HTML Editor

12pt Paragraph

0 words

Previous

Saving... Submit Quiz
