## Supplemental File and Appendix for "Designing a Clerkship Curriculum for Medical Students in Clinical and Medical Informatics in the Electronic Medical Record Era": A Survey of Regenstrief's Current Research - Pages Concatenated.docx

### Public Health Informatics

Core Faculty: Brian Dixon, Shaun Grannis, Chris Harle, Nir Menachemi and Joshua R Vest. This group works on developing theory in public health practice through the Fairbanks School of Public Health, as well as application in concert with the Marion County Health Department, Centers for Disease Control and Prevention and others.

Public health informatics generally seeks to:

apply health information and technologies to improve population health, including the surveillance and prevention of disease as well as general health promotion

This definition can otherwise be simply described as the intersection of technology and public health. Unfortunately, this negates the domain of informatics, and the definition promulgated by AMIA. Combining these two, we get:

the study and pursuit of the effective uses of biomedical data, information, and knowledge to improve population health, including the surveillance and prevention of disease as well as general health promotion

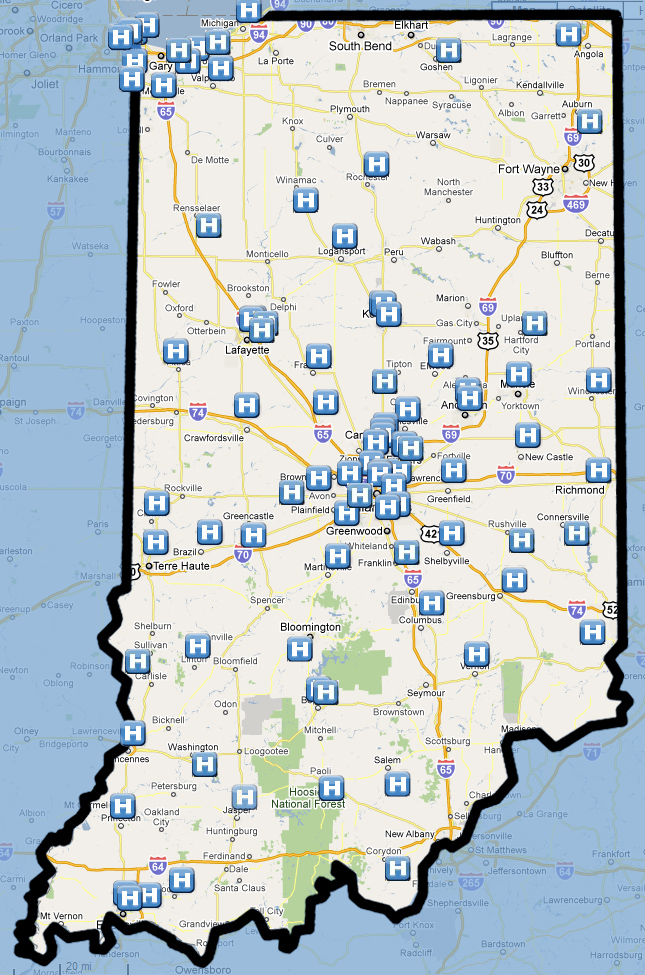

##### Patient Matching

A core expertise of the institute is in patient matching. As the United States lacks a national or state-level identifier capable of uniquely identifying a patient across multiple services, systems, times or locations, matching patients is a requirement for most electronic records. On the state level, the health information exchange, a system that links records from multiple facilities across the state, uses a global patient identifier to describe each unique patient. Indiana’s health information (Indiana Network for Patient Care or INPC) exchange extends through the entirety of the state. A map of the participating facilities follows.

Probabilistic methods such as that described by *Zhu et al* serve as the gold standard for record linkage. For such a task as newborn screening, public health officials can link vital recrods with newborn screening results within the INPC to identify patients who have not been screened. *Biro et al* shows how patient matching can occur beyond the medical record itself, linking census data with medical record data to answer questions surrounding such concepts as socioeconomic status. In short, much like how knowing how unique keys within a database are replicated within another, patient matching allows for the identification of unique patients across multiple databases, computer systems and more generally as a means to expand the data available on said patients.

Patient matching may also allow for the identification of patients with partial similarity, such as for recruitment in clinical trials. Identifying commonalities between patients to form a cohort on dimensions such as socioeconomic status, race, and pertinent clinical variables such as control of diabetes, could lead to recruitment in interventions designed to improve the health of diabetic populations.

### The Teaching Electronic Medical Record

Core Faculty: Blaine Takesue.

The teaching EMR (tEMR) is Indiana University School of Medicine’s contribution to the AMA Initiative to Improve Medical Education ([Creating the Medical School of the Future](https://www.ama-assn.org/ama/pub/about-ama/strategic-focus/accelerating-change-in-medical-education/innovations.page)). Details from the [IU Newsroom](http://news.medicine.iu.edu/releases/2015/10/teaching-electronic-medical-records-regenstrief.shtml) about this activity describe its early successes. Future work to incorporate the electronic medical record into medical education through the RIME framework is hoped to include the tEMR for students to pursue mastery of particular concepts.

### Global Health Informatics and OpenMRS

Core Faculty: Theresa Cullen, Burke Mamlin, Paul Biondich and Jonathan “JJ” Dick.

The global health program is supported by the [Academic Model Providing Access to Healthcare - Kenya Research Network](http://medicine.iu.edu/ampathresearch/) and the [Open Medical Record System](http://openmrs.org/) (OpenMRS), a community of open source electronic medical developers scattered around the world and based at Regenstrief. [AMPATH](http://www.ampathkenya.org/) is a collaboration between a [consortium](http://www.ampathkenya.org/our-partners/consortium-members/) of medical and global health institutions, from Indiana University, Brown University, University of Massachusetts, University of Toronto, Duke University, Icahn School of Medicine and University of California San Fransisco, to name a few.

##### Open Medical Record System

The OpenMRS system is used globally, with the [atlas](https://atlas.openmrs.org/) below.

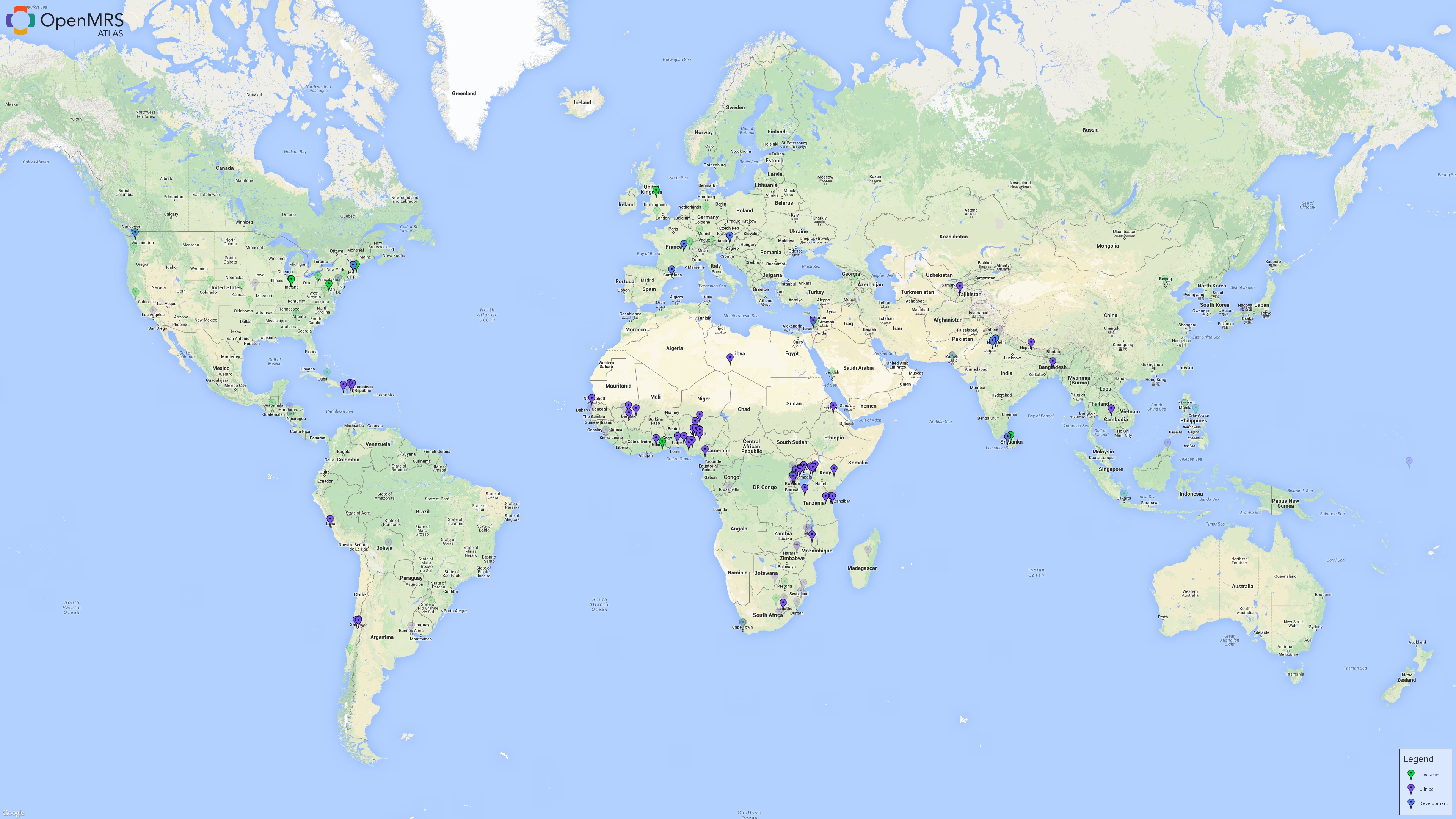

It serves as a means to enable electronic health record systems in low-resource settings. In addition, it is part of a global community of open source developers contributing to develop health information architecture. Another initiative is [OpenHIE](https://ohie.org/), an open source health information exchange platform that has allowed countries in Africa and elsewhere to effectively coordinate care across geographical and political entities.

### Hospital and Clinical Operations

Core Faculty: Paul Dexter, Jon Duke, Burke Mamlin, Titus Schleyer, JT Finnell, Justin Morea, Tim Imler.

Dr. Dexter, as the Chief Medical Informatics Officer at Ezkenazi Health, alongside his collaborators, contribute to the development of research activities that impact hospital practice. Primary areas that are currently under investigation include:

- Medication reconciliation
- Pharmacogenomics
- Data Analytics

This work is primarily driven by outcomes research and cohort definition through novel analytic platforms such as text mining, big data and health information exchanges.

### Pediatric Informatics

Pediatric informatics research is conducted both by scientists within and affiliated with the institute, as well as the Department of Pediatrics.

Core Faculty: Stephen Downs, Emily Webber, Paul Biondich, Aaron Carroll, William E Bennett, Marc Rosenman, and others.

##### Child Health Improvement through Computer Automation

Child Health Improvement through Computer Automation (CHICA) is a decision support system in the pediatric clinics at both Riley Hospital for Children and Ezkenazi Health. It works using the principles of expected utility to suggest questions to both pediatricians and patients that will help reduce the amount of overhead required for the pediatric encounter by guidelines and preventative screening. A comprehensive history of the CHICA system can be found in the 2015 article in Artificial Intelligence in Medicine by *Anand et al*. Further reading as to the complexities of the system and its operation can be found in the references below.

### Inpatient Pediatrics

On the inpatient side, the domain of pediatric health informatics differs from that of adults. To this end, Dr. Webber has done work to support the development of this unique subdiscipline, as well as support the translation of pediatric solutions to other facilities, both adult and pediatric. In addition to this work, Dr. Webber supports quality improvement activities in her role as the medical director for pediatric informatics at Indiana University Health and the subsequent leadership role that entails in the department of pediatrics section of hospital medicine, as well as Riley Hospital for Children. Dr. Webber also serves as the site coordinating faculty for the clinical informatics fellowship at IU Health.
