## Supplemental File and Appendix for "Designing a Clerkship Curriculum for Medical Students in Clinical and Medical Informatics in the Electronic Medical Record Era": Decision Science - Pages Concatenated.docx

### Fundamentals of Probability

Probability is a concept that describes the possible outcomes of a given event as a ratio between the possibility of an event’s outcome to all possible outcomes. A probability of **0** means that there is absolute certainty that an event will not occur. A probability of **1** means that there is absolute certainty that an event will occur. All other probabilities between **0** and **1** are graduated steps in between.

Several interpretations of the world through the lens of probability are often cited:

1. *Frequentist* philosophy suggests that sampling creates an understanding of the world: data are a repeatable random sample. Parameters are constant during this process.
2. *Bayesian* philosophy suggests that data observed from a sample is indeed from a realized sample. Parameters are described probabilistically, and therefore data is fixed.
3. *Classical* philosophy derives an expected value based on underlying mechanisms (such as the probability of a coin flip being **0.5** heads and **0.5** tails).
4. *Subjective* philosophy derives a probability based on subjective thought regarding the mechanisms and your knowledge of the past, combining classical and frequentist world views to present a measure of belief.

#### Biases in Estimating Probability

The seminal work exposing biases in the estimation of probability (and the use of decision science) is that of *Kahneman and Tversky*.

They describe several heuristics that modulate probability estimates, summarized below:

1. Representativeness: the estimation of the probability of an event based on how representative this event is of information provided, typically ignoring the logical considerations of the problem.
2. Availability: the estimation of the likelihood of an event based on ease of access to information.
3. Adjustment and Anchoring: the incorrect estimation of a probability, given the influence of a prior estimate, whether that estimate is adjusted, or used as a basis.

In addition to these empirically validated heuristics, another notion from social psychology is that of the vividness effect. *Taylor and Thompson*, in their work, describe this effect as a means by which elements of the availability heuristic are managed by our sense of “asthetic” or “vividness”, and a means by which elements of visual cognition have influenced cognitive models (as will be seen later).

- Kahneman, Daniel, and Amos Tversky. “Prospect theory: An analysis of decision under risk.” Econometrica: Journal of the Econometric Society (1979): 263-291.
- Tversky, Amos, and Daniel Kahneman. “Judgment under uncertainty: Heuristics and biases.” science 185, no. 4157 (1974): 1124-1131.
- Collins, Rebecca L., Shelley E. Taylor, Joanne V. Wood, and Suzanne C. Thompson. “The vividness effect: Elusive or illusory?.” Journal of Experimental Social Psychology 24, no. 1 (1988): 1-18.
- Taylor, Shelley E., and Suzanne C. Thompson. “Stalking the elusive” vividness" effect." Psychological review 89, no. 2 (1982): 155.

#### Bayes Theorem

Within the axioms that define the mathematical or logical rules of probability, there are several important definitions: Two events **A** and **B** are independent, if they do not have any interrelationships, and if they satisfy:

$$P\left( A\cap B \right)=P\left( A \right)P(B)$$

 If **A** and **B** are mutually exclusive, meaning that they do not occur at the same time.

$$P\left( A\cup B \right)=P\left( A \right)+P(B)$$

**Bayes Rule** states that given **A** and **B** are independent events:

$$\Pr\left( B | A \right)=\frac{Pr(B\cap A)}{Pr(A)}=\frac{\Pr\left( B \right)Pr(A|B)}{Pr(A)}$$

### Fundamentals of Information Theory and Encoding

Given a set of symbols for a fixed finite alphabet

$$S=\{s_{0},s_{1},\ldots,s_{K-1}\}$$

with probabilities

$$P\left( S=s_{K} \right)=p_{k}, k=0, 1, \ldots, K-1$$

These symbols serve as discrete quantized representations of content within an information source (such as a patient). Each symbol is considered statistically independent, and are emitted from a discrete memoryless source. This source fits into a communication channel, as illustrated in the figure below.

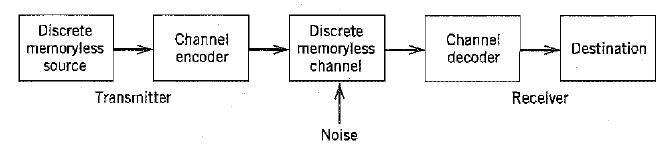

This channel serves to link source and destination via blocks, where each block consists of **n** successive source symbols.

*Blois*, in his book *Information and medicine: the nature of medical descriptions*, describes that the diagnostic process as the encoding of a series of signs and symptoms as a disease object. The fundamentals of probability serve as a means to describe the probability of events, as well as mathematically combine symbols emitted by information sources.

The amount of *information* gained after observing the event $S=s_{k}$ is quantified by the equation

$$I\left( s_{k} \right)=\log_{q} (\frac{1}{p_{k}})$$

Where qq is the number of symbols in the source alphabet. *Entropy* is defined as the average information content per source symbol

$$H\left( S \right)= \sum_{k=0}^{K-1} p_{k}I(s_{k})$$

Quantifying disease using the information and entropy allows for the representation of the probabilistic linkages between signs and symptoms.

Clinical data is encoded by terminology standards, as well as other modalities, in a science termed by *Feinstein* as *Clinimetrics*. Encoding information through quantization, scales and other forms allows for the same benefit as analog to digital conversion. This benefit comes in term of the ability to measure information content, perform symbolic logic operations, and in turn embrace a more quantitative view of clinical practice. This can be seen in the work of *Feinstein* and others in describing statistical theories of medicine.

- Feinstein, A. R. (1987). Clinimetrics. Yale University Press.
- Blois, Marsden S. *I1nformation and medicine: the nature of medical descriptions*. University of California Press, 1984.

### Shannon, Weiner and Cybernetics

*Claude Shannon* is argued to be the originator of the concept of information theory, beginning with his seminal thesis *A symbolic analysis of relay and switching circuits*, published both as an MIT thesis, and adapted later for the Transactions of the American Institute of Electrical Engineers. *Norbert Wiener* was a contemporary of Shannon, and served as a major driving force behind the notion of cybernetics, and a driving personality behind a great deal of post-war multidisciplinary science. Building on the notion of information theory, cybernetics is a means by which interrelations between elements within a system can be described. It is the main core of science from and during the 1950s that led to the notion of interdisciplinary as a core concept in modern science. As described in Weiner’s *Cybernetics or Control and Communication in the Animal and the Machine,* **cybernetics** is the science of control and communication in the animal and machine, typically reflected in the inclusion of control theory as a means to describe the relationship between information and action. This theory was influenced by a second degree relationship to Walter B Cannon, who coined the term **homeostasis** at Harvard/MIT. Furthermore, given an understanding of the infomedical model of disease, we have a passing familiarity with the notion of cybernetics. In the infomedical model of disease, information sources from each element of the model (society, mind, body, gene, biosphere and person) complete control loops intended to inform the diagnostician and be altered by subsequent evaluation… All as a means to uncover a more accurate model of disease and its interactions with other elements of the model and in turn, world. Norbert Wiener’s 1952 Alvarenga Prize lecture to the College of Physicians of Philadelphia, entited “The Concept of Homeostasis in Medicine” further demonstrates and explores the most common application of cybernetics to medicine, as taught in the basic science (physiology) curriculum: that of homeostasis.

Building on the notions of information theory and cybernetics, *McCulloch and Pitts* presented their *logical calculus of the ideas immanent in nervous activity* as a means of describing the functioning of the brain and neurons, and their idea for the linkages between neural physiology and decision-making. The life of Walter Pitts and the circumstances by which his scientific endavours occurred was recently detailed in a piece in the Nautilus magazine, entitled [The Man Who Tried to Redeem the World with Logic](http://nautil.us/issue/21/information/the-man-who-tried-to-redeem-the-world-with-logic).

- Wiener, N. (1949). Cybernetics or Control and Communication in the Animal and the Machine. 6th Printing, The Technology Press.
- McCulloch, W. S., & Pitts, W. (1943). A logical calculus of the ideas immanent in nervous activity. The bulletin of mathematical biophysics, 5(4), 115-133.
- Shannon, C. E. (1938). A symbolic analysis of relay and switching circuits. American Institute of Electrical Engineers, Transactions of the, 57(12), 713-723.
- Shannon, C. E. (1937). A Symbolic Analysis of Relay and Switching Circuits (Masters dissertation, Massachusetts Institute of Technology).
- Wiener, Norbert. “The concept of homeostasis in medicine.” Transactions & studies of the College of Physicians of Philadelphia 20, no. 3 (1953): 87-93.

### Linking Information Theory and Probability: The Bayesian Cognitive Model

Given an information theoretic model that accepts probabilistic representations; symbols, alphabets and language converge as the link between information theoretic approaches to modeling communication and Bayesian probability. *Tenenbaum et al* and *Clark* describe the notion that inferences are a key component of concepts such as: acquiring language, pattern recognition and grasping causal relations. These inferences are described in probabilistic terms as having Bayesian character. From this, *Clark* and *Tenenbaum et al* describe the notion that this character informs our current understanding of the notion of how the brain communicates internally, as well as with external actors. As such, Bayesian probability, and information theoretic encoding methods allow for the performance of tasks such as the aforementioned: acquiring language, pattern recognition and grasping causal relations. Whether or not this fits within current views on cognitive science as a means to describe the world is argued for in *Evans and Patel*’s work describing the notion of:

the science of human information processing, concerned with how people learn , remember and solve problems and as such might be thought of as a ‘basic science’ of education.

They suggest a reformation of the medical education and practice curriculum to fit within their models of the world, much as cognitive psychologists have dominated the field of education. Whether this is correct, many institutions have gone down this pathway, through both problem-based learning in the case of *Barrows*, or “systems medicine” in the case of *Sturmberg et al*, and current methods of medical reform are directed towards a combination of the two (through the four pillars approach and beyond).

- Tenenbaum, J. B., Kemp, C., Griffiths, T. L., & Goodman, N. D. (2011). How to grow a mind: Statistics, structure, and abstraction. Science, 331(6022), 1279-1285.
- Clark, A. (2013). Whatever next? Predictive brains, situated agents, and the future of cognitive science. The Behavioral and Brain Sciences, 36(3), 181-204.
- Evans, D., & Patel, V. (1989). Cognitive Science in Medicine: Biomedical Modeling. MIT Press.
- Barrows, H. S. (1980). Problem-based learning: An approach to medical education. Springer Publishing Company.
- Sturmberg, J. P., Martin, C. M., & Katerndahl, D. A. (2014). Systems and Complexity Thinking in the General Practice Literature: An Integrative, Historical Narrative Review. The Annals of Family Medicine, 12(1), 66–74.
- Pock, A. R., Pangaro, L. N., & Gilliland, W. R. (2015). The “Pillars” of Curriculum Reform. Academic Medicine.

### Fundamentals of Decision Science

Decision science is a discipline that combines cognitive science and economics, builds on the work of *Von Neumann and Morgenstern*. It allows for the description of rational behavior of systems of choice (economic systems), incorporating notions of human decision making from the psychology literature and practice. *Detsky, Naglie, Krahn, Redelmeier and Naimark* produced a primer series for the interested reader that describes decision analysis in more detail.

- Von Neumann, J., & Morgenstern, O. (2007). Theory of games and economic behavior (60th Anniv.). Princeton university press.
- Detsky AS, Naglie G, Krahn MD, Naimark D, Redelmeier DA. Primer on medical decision analysis: Part 1—Getting started. Medical Decision Making. 1997 Apr 1;17(2):123-5.
- Detsky AS, Naglie G, Krahn MD, Redelmeier DA, Naimark D. Primer on medical decision analysis: part 2—building a tree. Medical decision making. 1997 Apr 1;17(2):126-35.
- Naglie G, Krahn MD, Naimark D, Redelmeier DA, Detsky AS. Primer on medical decision analysis: part 3—estimating probabilities and utilities. Medical Decision Making. 1997 Apr 1;17(2):136-41.
- Krahn MD, Naglie G, Naimark D, Redelmeier DA, Detsky AS. Primer on medical decision analysis: Part 4-Analyzing the model and interpreting the results. Medical decision making. 1997 Apr 1;17(2):142-51.
- Naimark D, Krahn MD, Naglie G, Redelmeier DA, Detsky AS. Primer on medical decision analysis: part 5—working with Markov processes. Medical decision making. 1997 Apr 1;17(2):152-9.

#### The Discrete Choice

Economics, or the study of choice, relies on the notion that choices are discrete in nature. As such, decision science builds on this framework to describe the required three components of a decision:

1. The decision has two or more options as to courses of action. An example of how these courses can be represented is in a decision tree.
2. There is uncertainty as to which of those options have a preferential outcome. This uncertainty has a representative model in probability.
3. These outcomes are capable of eliciting preferences. These preferences are typically expressed by utilities.

The concept of utility can be broadly described as the quantification of a preference for an outcome. *Drummond et al* describe utility in the context the notions of utility, value and preference as components of measurement. Specifically, along the axes of response methods and question framing:

Question Framing

Response Method

Certainty (values)

Uncertainty (utilites)

Scaling

Rating scale, category scaling, visual analogue scale, ratio scale

Choice

Time trade-off, paired comparison, equivalence, person trade-off

Standard gamble

Specifically, the notion that a question framed under uncertainty (in that the choice has a probability) can be described as one that would capture a “utility” towards a choice or scaled value. This utility can be described in this sense as a “preference adjusted for risk”, where risk is expressed as the subject’s attitude towards the choice in question (such as that elicited through the standard gamble).

Decisions can be modeled by the use of a decision tree, a representation of branching decisions starting from a point of information (decision node) and branching on each choice with a probability (chance node) to end at a terminal decision point (terminal node). An example of a decision tree describing the choices of observation or surgery leading to death or survival. In this case, the terminal nodes describe simple utilites of either 1 (survival) and 0 (death).

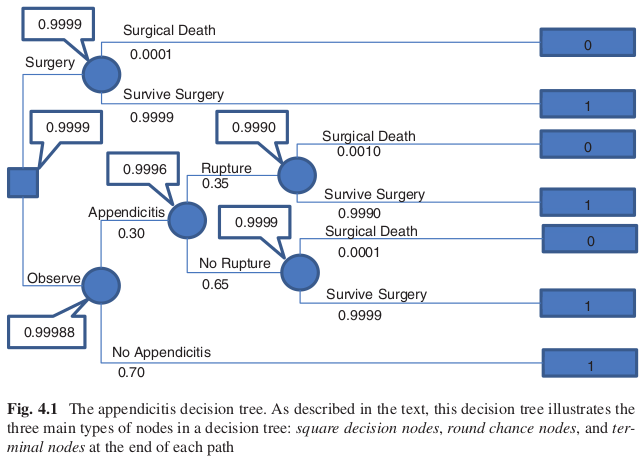

- Drummond, M. F., Sculpher, M. J., Torrence, G. W., O’Brien, B. J., & Stoddart, G. L. (2005). Methods for Economic Evaluation of Health Care Programmes (Third.). Oxford University Press.

#### Expectation

Expected value theory suggests that for an event (**E**) the expected value of that event is the multiplication of the value of the event and the probability of the event occurring plus the naught of the event and its value.

$$EV\left( E \right)=v\left( \neg E \right)\Pr\left( \neg E \right)+v\left( E \right)Pr(E)$$

The notion of an expectation is extended into the utility space. Specifically, it serves to extend expected value theory into the preference space. This results in the following formula:

$$EU\left( E \right)=u\left( \neg E \right)\Pr\left( \neg E \right)+u\left( E \right)Pr(E)$$

For a choice system, whether expressed as a decision tree or as a chain of events, the expected value of a choic can be calcualted for each branch of the decision node by using the expectation formula. Utility allows for a more appropriate scaling of the events than the simple value function, as described by prospect theory (decision makers are more sensitive to losses than gains). Furthermore, expected utility theory proffers a set of axioms underlying its functional form and application described in detail by *Howard*:

1. Orderability
2. Transitivity
3. Monotonicity
4. Decomposability
5. Continuity and Substitutability

- Kahneman, D., & Tversky, A. (1979). Prospect Theory: An Analysis of Decision under Risk. Econometrica, 47(2), pp. 263–292.
- Howard RA. Risk preference. Readings on the principles and applications of decision analysis, vol. II. Menlo Park: Strategic Decisions Group; 1983.

#### Benefits and Costs

In addition to utility, decisions can be weighted by other forms of measurement. In particular, two are typically used in decision analysis: benefits and costs. The former serves to characterize clinical outcomes and metrics. The latter allows for the accounting of monetary assets. In order to function in the formulae in a similar manner to utilities, the core requirement is that they be unit-defined.

Particularly with clinical outcomes, unit-defined measurement is often difficult. One example of a standard metric (to which outcomes are often mapped onto) is that of the Quality Adjusted Life Year (QALY). This serves as a measure of the impact that a particular clinical action has on the morbidity and mortality of the patient.

#### Markov Models - The Finite State Machine

Beyond the decision tree is the more complex process of the economic markov model. This model operates similar to a finite state machine, operationalizing the transition of a signal (or in this case a patient) from state to state. These states describe the disorder and context of care, and through simulation we can better understand population behaviour.

### The Application of Decision Science

With theory in hand, one can begin to perform evaluations on technology and clinical procedures. The [WHO](http://www.who.int/medical_devices/assessment/en/) describes Health Technology Assessment (HTA) as:

the the systematic evaluation of properties, effects, and/or impacts of health technology. It is a multidisciplinary process to evaluate the social, economic, organizational and ethical issues of a health intervention or health technology. The main purpose of conducting an assessment is to inform a policy decision making.

The element of HTA that serves to understand the decisions from a cost, utility, and benefit perspective is a prominent application of decision analysis. The Bayesian cognitive model and its precurors allow for more accurate descriptions of decision-making, both the processes at play in performing the decision-making act and the disease itself.

### Assignment: Modeling a Decision Problem

### Modeling a Decision Problem

1. Select your favourite clinical decision or circumstance. Write out a problem definition.
2. Diagram this problem using your preferred method (such as dot) to create an influence diagram or decision tree (dot documentation can be found [here](http://www.graphviz.org/Documentation.php)).
3. Describe how you would evaluate the decision using cost, benefit or utility.
4. Perform a back of the envelope calculation that will help you assess the best solution to your clinical problem.
