## Supplemental File and Appendix for "Designing a Clerkship Curriculum for Medical Students in Clinical and Medical Informatics in the Electronic Medical Record Era": Final Project - Pages Concatenated.docx

### Introduction

Your task is to write a design document detailing a decision support system for your primary specialty of interest, building upon a clinical guideline/literature. Specifically this task includes:

1. Writing a design document describing the clinical decision you wish to address, and the clinical problem it revolves around.
2. Design a database to describe the information sources capable of feeding into such a designed solution.
   - Given the [OpenMRS Data Model](https://wiki.openmrs.org/display/docs/Data+Model), describe what data is required by your application
   - If you have completed the requisite mini-course, Use Unified Modeling Language (UML) to detail the functional needs of your project, given objects from the OpenMRS data model
3. Write in Python (or pseudo-code) the code necessary to access your described database/data model. This code should provide the clinical logic at point of care that would accompany the system you have designed in the hospital/clinical environment you have defined.
4. Write a final report, detailing as much of the solution as possible
   - Detail the design of your decision support system, as well as the rationale for each component given the clinical circumstances
   - Ensure you describe the clinical circumstances of the problem, and ponder hypotheses that this solution would be capable of influencing or testing This report should integrate the knowledge you have gained throughout this course, specifically that from other modules to supplement your design document in a final design.

### What is a Design Document?

#### Where to Start?

Using [UpToDate](http://www.uptodate.com/contents/search), you can search for disease topics (or treatment strategies, tests…) and from the results, form a basis to understand the context of that disease’s problem, computerization, guidelines and other circumstances. This will afford you a basic tool to start framing the problem definition, however, it should not be used in isolation. Searching the medical literature through PubMed, Google Scholar or other services will be helpful. In addition, you can specify the information detailed by a search via UpToDate, such as searching solely for the differential, interpretation or child-based context of a disease or test. This specificity may help you further detail the specifications and requirements of your design solution.

#### Problem Definition

A problem definition is somewhat akin to a hypothesis, in that it states the problem to which the design is intended to address (an analogy would be describing the question this design intends to solve). The problem definition must describe the problem as to provide a context for the solution, as well as the basis from which requirements and specifications can be drawn. To build the problem definition, it is suggested that you start off with a short single statement, and build from there to flesh out the problem definition.

Examining the literature on the identification and evaluation of children with autism spectrum disorders, we can see that the screening could be improved by automatic calculation of age, and patient-facing data collection.

#### Requirements

This section describes the requirements that must be fulfilled for the design to adequately address the problem definition (you can think of it as an answer to the question posed in the problem definition). In describing the requirements, you must indicate as to how these are gleaned from both the problem definition and context of the problem itself.

We require the design to:

1. Have the most amount of automation to collect information from patients and compute scores/automate passthrough of the algorithm.
2. Provide decision support as to the propogation through the algorithm (both end-stage, as well as automated decisions made)

#### Specifications

These are the elements of the design necessary to fulfill the requirements (you can think of this as the rationale for the requirements). Importantly, this section must cover both the requirements as well as link back to the problem definition. This should result in a technical description of the design in question, and allow for the easy transition from design document to prototype.

The design shall:

1. Prompt the parent for answers the questions:
   1. Does the patient have a sibling with ASD
   2. Do you as a parent have a concern regarding ASD and your child’s development
   3. Have any other caregivers expressed concern regarding ASD and your child’s development
2. Prompt the pediatrician for an expression of concern regarding ASD
3. Score the questions in (1) and (2) to identify where to progress, and from there, inquire after the age of the patient.
4. Progress through the algorithm, prompt the pediatrician for the classification of the results of appropriate evaluations/screening tools and hit the end stage (7a), (7b), (8) of the algorithm.
5. Report the algorithm’s progress and decisions.

#### Prototype/Functional Design Solution

This section articulates the prototype/solution in detail, both describing and implementing the design. Descriptive languages such as VHSIC Hardware Description Language, hardware computer assisted design layouts, computer code, mechanical drawings,… are examples of information in this section.

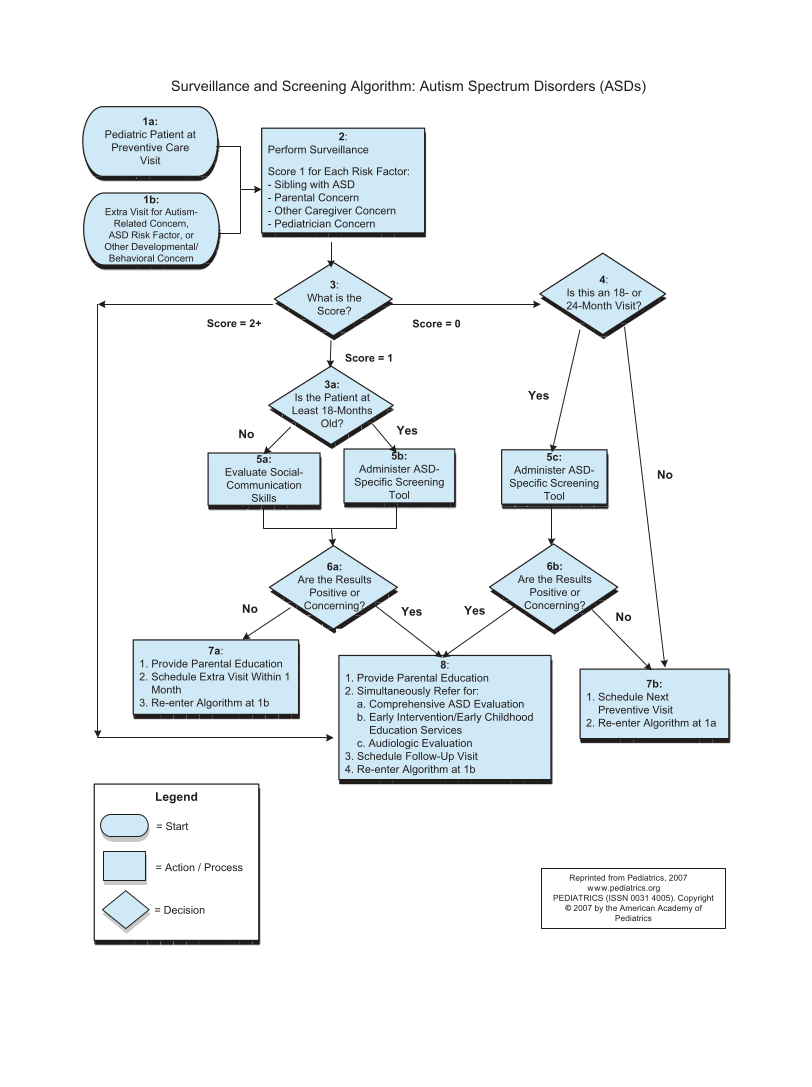

- Johnson, C. P., & Myers, S. M. (2007). Identification and evaluation of children with autism spectrum disorders. Pediatrics, 120(5), 1183-1215

### Assignment: Design Document Deliverable

Write a design document describing the clinical decision you wish to address, and the clinical problem it revolves around.

### Assignment: Computer Logic Deliverable

Write in Python (or pseudo-code) the code necessary to access your described database/data model. This code should provide the clinical logic at point of care that would accompany the system you have designed in the hospital/clinical environment you have defined.

### Assignment: Database Schema Deliverable

 Design a database to describe the information sources capable of feeding into such a designed solution.

1. Given the [OpenMRS Data Model](https://wiki.openmrs.org/display/docs/Data+Model), describe what data is required by your application
2. If you have completed the requisite mini-course, Use Unified Modeling Language (UML) to detail the functional needs of your project, given objects from the OpenMRS data model

### Assignment: Final Report Deliverable

Write a final report, detailing as much of the solution as possible

1. Detail the design of your decision support system, as well as the rationale for each component given the clinical circumstances
2. Ensure you describe the clinical circumstances of the problem, and ponder hypotheses that this solution would be capable of influencing or testing

   This report should integrate the knowledge you have gained throughout this course, specifically that from other modules to supplement your design document in a final design.
