## Supplemental File and Appendix for "Designing a Clerkship Curriculum for Medical Students in Clinical and Medical Informatics in the Electronic Medical Record Era": Fundamentals of Medical Informatics - Pages Concatenated.docx

### Definition of Disease

Beginning in 1643 with his seminal *Principia Philosophiae*, *René Descartes* significantly influenced science by contributing the thought that both mind-body union and separation are opposing concepts. Subsequently, this led him to commit to the course of mind-body dualism, the root of which in [modal metaphysics](http://plato.stanford.edu/entries/descartes-modal/) and its influence on philosophical & scientific thought can be seen through the [InPhO project](https://inpho.cogs.indiana.edu/). Mind-body dualism and the philosophical ideas of Descartes, subsequently led to the theory that the biomedical and psychological elements of disease (and bodily function) were significantly distinct and separate entities.

This idea dominated science and the practice of medicine for the next few hundred years, and led to the coining of the *biomedical model of disease*, in which disease is solely a function of molecular biology and biochemistry. From this, disease can be described as: *observed dysfunction of molecular and physiological processes as seen from the measurements which the physical and natural sciences can conduct on the body*.

### The Biopsychosocial Model

In the mid 1970s, *Fabrega* and *Engel* introduced the notion of integrating the biological and social sciences, building on the modern concept of the *study of medical systems*. This integration resulted in the *biopsychosocial model of disease*. This concept does not reconcile mind-body dualism rather it builds upon the biomedical model, and suggests that the concept of disease should include both biological and psychosocial facets. Included in this new definition was the notion that the biomedical model was of reductionist character. In keeping with the post-war progression of science from a reductionist to a holistic or complex mindset, it further invoked the notion of general systems theory (hinting at the concept of the study of medical systems) as a means of describing the reconciliation of biomedical and psychosocial concepts.

- Engel, G. L. (1977). The Need for a New Medical Model: A Challenge for Biomedicine. Science, 196(4286), pp.129-136.
- Engel, G. L. (1981). The clinical application of the biopsychosocial model. Journal of Medicine and Philosophy, 6(2), 101-124.
- Fabrega Horacio, J. (1975a). The Need for an Ethnomedical Science. Science, 189(4207), pp.969-975.
- Fabrega Horacio, J. (1975b). The position of psychiatry in the understanding of human disease. Archives of General Psychiatry, 32(12), 1500-1512.

### The Infomedical Model

Beginning with *Foss and Rothenberg*, the information age of biomedical science has suggested a further advancement: *the infomedical model of disease*. This model more aptly interacts with advances in bioinformatics and modern theories in biomedical sciences promulgated by *Brenner*. It suggests that the model of disease is indeed a complex system and has multiple axes (society, gene, mind, body, biosphere). This model is visually detailed in the frontispiece of *Foss and Rothenberg* (duplicated below).

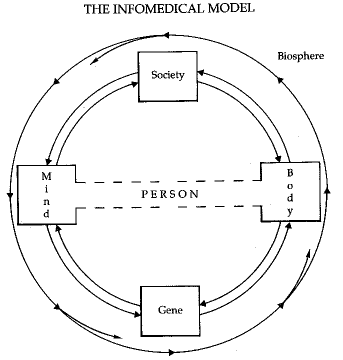

The infomedicine model, and the biopsychosocial model preceding it, supports information theory and complex systems science in their roles as methods capable of synthesizing knowledge from multiple sources and applying it to care in an iterative, controlled process. This is particularly pertinent given the National Research Council’s report from 2011, entitled *Toward Precision Medicine: Building a Knowledge Network for Biomedical Resarch and a New Taxonomy of Disease*. This report suggests that moving forward, the strategy of biomedical research should be to push forward a new taxonomy of disease based on molecular biology (and advances thereof). Such a putsch against the current science is an attempt to forcibly integrate concepts related to the feedback system illustrated by the infomedical model into the practice of science, exemplified by the translational medicine process, as the core of scientific activity. Translational medicine, as described by *Woolf*, comes from the Institute of Medicine’s [Clinical Research Roundtable](http://iom.nationalacademies.org/Activities/Research/ClinicalResRT). This roundtable specifically defined two core phases:

### T1

the transfer of new understandings of disease mechanisms gained in the laboratory into the development of new methods for diagnosis, therapy, and prevention and their first testing in humans

### T2

the translation of results from clinical studies into everyday clinical practice and health decision making

Other phases include T3 and T4, which expand translational science into the practice implementation and policy phases. During these phases, science is tested in a pragmatic or real-world setting and its impact on society through policy and high-level engagement evaluated. Translational science and the paradigm which it has influenced within biomedical science, has the potential to influence medicine in the years to come given the 2012 establishment of the [National Center for Advancing Translational Sciences](https://ncats.nih.gov/).

- Foss, L., & Rothenberg, K. (1987). The Second Medical Revolution: From Biomedical to Infomedical. Boston: New Science Library.
- Brenner, S. (2012a). History of science. The revolution in the life sciences. Science (New York, N.Y.), 338(6113), 1427–8.
- Brenner, S. (2012b). Turing centenary: Life’s code script. Nature, 482(7386), 461–461.
- National Research Council (US) Committee on A Framework for Developing a New Taxonomy of Disease. (2011). Toward precision medicine: Building a knowledge network for biomedical research and a new taxonomy of disease. National Academies Press (US).
- Woolf, S. H. (2008). The meaning of translational research and why it matters. JAMA, 299 (2), 211-213.

### Historical Perspectives

The question of **How does one practice medicine?**, and whether medicine adheres strictly to the tenets of scientific inquiry or whether there truly is an art to medicine, has been debated since before *Sir William Osler* and *Abraham Flexner* reformed American medical education.

*Guthrie* details a path by which the Scottish medical education establishment took root in the United States, influencing both the granting of the degree *Medicinae Doctor* akin to the Ancient universities of Scotland (Aberdeen, Edinburgh, Glasgow, and St. Andrews) as well as the prominence of certain academic sources of medical education prior to the Flexner report. *Ludmerer* provides more detail as to the evolution of medical education in the United States, and details changes in medical education and their influence on medical practice. As *Guthrie* mentions; given the time, while *Flexner* advocated for a more focused biomedical education (modeled after the curriculum of German laboratory sciences and medical schools), *Osler* suggested that the introduction of classical education and the humanities to medical education served exemplary value. Both of these men suggest a depth of practice that takes into account the nature of medical education as well as the founding principles of the (medieval) university. To detail these principles, both *Flexner* in his *Universities* and *Osler* in his *Aequanimitas* cite the eminent Oxonian, Cardinal John Henry Newman:

A University consists, and has ever consisted, in demand and supply, in wants which it alone can satisfy and which it does satisfy, in the communication of knowledge, and the relation and bond which exists between the teacher and the taught. Its constituting, animating principle is this moral attraction of one class of persons to another ; which is prior in its nature, nay commonly in its history, to any other tie whatever; so that, where this is wanting, a University is alive only in name, and has lost its true essence, whatever be the advantages, whether of position or of affluence, with which the civil power or private benefactors contrive to encircle it.

How the above aligns with further advancements in the United States can be seen in further literature, and in the broad strokes of the writing of*Ludmerer*.

Although historical perspectives provide us with a sense of how thinking has evolved and the knowledge contained within the medical degree, the definition of what it is to practice of medicine remains. *Osler* in his book entitled *The Principles and Practice of Medicine: Designed for the Use of Practitioners and Students of Medicine*, quotes Plato’s *Gorgias*:

And I said of medicine, that this is an art which considers the constitution of the patient, and has principles of action and reasons in each case

Further, *Cannon*, in his treatise *The Use of Clinic Records in Teaching Medicine* describes the physician work as summarizing concisely the*work of the physician*. Building upon that sentiment, he describes that the physician’s work has two facets:

1. The observation of data
2. The rational judgement of data

*Cannon* and *Osler* both allude to the core concept in medical practice being that of diagnosis. *Sir Thomas Clifford Allbutt* in the preface to the first edition of his seminal work *A System of Medicine*, describes the role of the physician as a *thoughtful man versed in natural inquiry*, seeking to raise a patient up to the highest possible health (at which perfect health in an abstract infinite state) as an *engineer who cannot construct, but is skilled in conservation and repair*. This task belies the central notion of understanding the system, as diagnosis (or in the analogous profession of engineering, the translation of requirements to specifications through inquiry) is essential to the analogized practice. Furthermore, in examining the notion of diagnosis and it as in turn a core element of the practice of medicine, he views curative action as an art, the application of sciences and the concern of the physician. As he further delves into nosology (important given his book’s status as one of the earliest compendiums/nosologies of concepts in medicine), *Allbutt* describes clinical diagnosis as:

not investigation - a distincition some practitioners forget; diagnosis depends not upon all facts, but upon crucial facts. Indeed we may go farther and say that accumulation of facts is not science; science is our conception of the facts: the act of judgment, perhaps of imagination, by which we connect the unknown with the known.

Describing the theory of medical practice in turn with the definition of disease allows us to understand the information sources inherent to the practice of medicine. To assist in this goal, the next two sections describe two facets of theory centring on the core concept in medical practice: that of diagnosis.

- Osler, W. (1919). The old humanities and the new science: The presidential address delivered before the Classical Association at Oxford, May, 1919. British Medical Journal, 2(3053), 1. (A)
- Flexner, A. (1910). Medical education in United States and Canada: a report to the Carnegie Foundation for the Advancement of Teaching (No. 4). Carnegie Foundation for the Advancement of Teaching. (B)
- Wallis, F. (2000). Signs and Senses: Diagnosis and Prognosis in Early Medieval Pulse and Urine Texts. Social History of Medicine, 13(2), 265–278. (A)
- Ludmerer, K. M. (1999). Time to heal: American medical education from the turn of the century to the era of managed care. Oxford University Press. (B)
- Guthrie, D. (1959). The influence of the Leyden School upon Scottish medicine. Medical history, 3(02), 108-122. (A)
- Flexner, A. (1994). Universities: American, English, German. Oxford University Press. (B)
- Osler, W. (1922). Aequanimitas: With Other Addresses to Medical Students, Nurses and Practitioners of Medicine. P. Blakiston.
- Osler, W. (1910). The principles and practice of medicine: designed for the use of practitioners and students of medicine. D. Appleton. (B)
- Cannon, W. B. (1901). The Use of Clinic Records in Teaching Medicine. Bulletin of the American Academy of Medicine, 5(4), 203-213.
- Allbutt, T. C. (Ed.). (1896). A system of medicine (Vol. I). Macmillan and Co..

### Linguistic Theory

*Wallis* introduces a character of medieval medicine, from the abbey of Monte Cassino in central Italy of that dismisses the semiotics of clinical diagnosis and prognosis. Semiotics, driven by the American philosopher *Charles S Peirce*, is the linkage described by both *Burnum* and *Nessa*as the link between, signs & symptoms, disease and diagnosis. Visually, this can be seen in the following figure from *Nessa* (duplicated below).

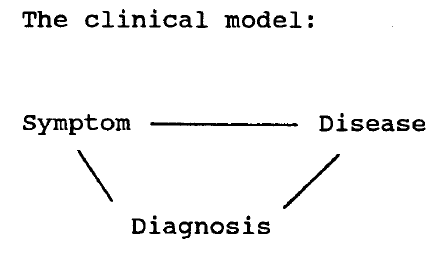

Philosophically, semiotics offers a means by which the linguistics of patient observation links with the categorization of disease via diagnosis. This method, although undoubtedly influenced by *Ferdinand de Saussure*, is still taught in medical coursework at Perugia and Padova. Semiotics provides a roadmap for the logical link between signs & symptoms and disease, and a concrete place for the art of diagnosis within the practise of medicine. While scientific fact and theory may support the link between signs & symptoms, it offers an empirical mechanism for diagnosis to have meaning within the context of scientific observation and the theory of disease.

Within the practice of medicine, the difference between science and arts, analogous to, in disease, the difference between mind and body, or biomedical and psychosocial, offers us a starting point for framing the disconnect solved by semiotic theory and reinforced by statistical empiricism. The modern demarcation between the arts and sciences in undergraduate education further complicates this problem, with the traditions of the (medieval) university and intellectual learning required for the course of the undergraduate degree upended by a focus on specialized, vocational and technical education. Rather than looking to fulfill the breadth of requirements instilled by the trivium and quadrivium into the basis of the bachelor’s degree and in turn the requirements for medical school, *Sommet et al* describes that we now have a focused*numerus clausus* based education system which fails to educate physicians. *Buchanan*, taking a cue from *Osler*, argues that medicine, in particular the clinical and laboratory arts, is a unique *strictly medical science*, differing from other elements of the liberal arts, but also that of the sciences. This precepts a theory of medicine, reinforced by the notion of biomedical ethics; the current practice of philosophy as applied to medicine. Buchanan builds on semiotic theory, and posits that the Galenic philosophy of signs and symptoms linked to disease requires meaning and can be in turn, satisfied by the liberal arts. These liberal arts, rather than being a distorted discipline that they are today, are instead the medieval trivium and quadrivium:

1. Grammar
2. Rhetoric
3. Logic
4. Arithmetic
5. Geometry
6. Music
7. Astronomy

Unlike the arts and humanities discussed in colleges today, these liberal arts serve as mechanisms for describing the world in a fashion seen prior to Flexner and the emphasis on laboratory sciences, to which Maycock, in his introduction to *Buchanan* attributes the death of the clinical arts. This notion of the clinical arts as a component of medicine and the influence language and semiotics have on the practice of medicine, is furthered by *Crookshank* in his supplementary essay in *A Study of The Influence of Language upon Thought and of the Science of Symbolism*.*Crookshank* suggests that language and semiotics are important in the study of medicine, predominantly to fill the void left by solely scientific fact and theory. The lack of the principles of a theory of medicine, as alluded to in *Buchanan*, prevent Crookshank from referring to it as solely a science, rather, he suggests that scientific theory and empirical information supports the practice of medicine and its role in diagnosing and treating disease. Crookshank further alludes to the lack of a scientific theory of disease as described by *Allbutt* and others, and rather describes disease as a classification scheme inherently built on some form of linguistics, with scientific relations and facts used in the descriptive. While this holds in some sense leading into the modern ontological era, via the circumstances by which we retain nomenclature and disease relationships from earlier times, as well as the relationships between disease and forms not explicitly described by scientific theory and fact; practice in psychiatry and somatic medicine alludes strongly to the existence of the permeance of linguistic ideas within the concepts of disease and diagnosis. Taking forward the theory of medicine described by *Buchanan*, supported by the philosophical and linguistic arguments of semiotics, we can describe the practice of medicine as the inference of meaning from signs and symptoms which in turn are related to disease, and the subsequent translation of that meaning into an actionable course of treatment. Such treatment, inference and logic is informed primarily by scientific fact and theory, and supported by statistical and mathematical logic. With this in mind, statistical mechanisms serve to shore up the holes left in the structures and definitions of disease as they are developed, and semiotics and linguistics lay out a foundation for practice and logic, extended in turn by mathematical calculation and empiricism.

- Buchanan, S. (1938). The Doctrine of Signatures A Defense of Theory in Medicine. University of Illinois Press.
- Sommet, N., Pulfrey, C., & Butera, F. (2013). Did my MD really go to university to learn? Detrimental effects of numerus clausus on self-efficacy, mastery goals and learning. (A)
- Nessa, J. (1996). About signs and symptoms: Can semiotics expand the view of clinical medicine? Theoretical Medicine, 17(4), 363–377.
- Burnum, J. F. (1993). Medical Diagnosis through Semiotics: Giving Meaning to the Sign. Annals of Internal Medicine, 119(9), 939–943.
- Malinowski, B., Crookshank, F. G., Ogden, C. K., &amp; Richards, I. A. (1927). The Meaning of Meaning; a Study of the Influence of Language Upon Thought and of the Science of Symbolism. Harcourt, Brace.

### Statistical Theory

Beginning with *Ledley and Lusted*, decision science has promulgated a theory of diagnostic reasoning and computationally supported clinical judgement that is the basis behind a great deal of diagnostic research today. *Feinstein*, in a series of publications, explored the concept of clinical algorithms and decision making, alongside the integration of biomedical knowledge. He describes this approach as being

Based on mathematical “grammar” rather than clinical “idioms,” these new diagnostic approaches bear no resemblance to conventional forms of clinical reasoning. The new mathematical proposals for diagnostic decisions are often derived from a concept of probability called Bayes Theorem.

These methods differ from the linguistic theory which suggests linkages between concepts and described information, rather than statistical formalisms representing such linkages. *Feinstein*, in his perspective piece in the Annals of Internal Medicine, offers commentary on the distraction of the application quantitative models (from fields such as decision analysis and psychometry), rather than a focus on *data, taxonomy and reasoning*. Such problems are indicative of the further research in following years (given that this article was published over 20 years ago) in medical informatics; focusing on data and clinical nosology/taxonomy.

*Norusis and Jacquez* echo Feinstein, formalizing the mathematical linkages between symptoms and disease, and describing in mathematical terms elements of the diagnostic discrimination process. *Szolovits and Pauker* further describe the use of computers to deploy such decision science and statistical frameworks. These examples are early methods by which statistical means can be applied to solve diagnostic logic. This logic, when put into practice, results in physicians having an understanding of the risk behind the differential diagnosis in question, as it applies to the patient they are investigating. In turn, this risk, and its application in practice is subject to bias and error. *Croskerry* describes the decision sciences behind a statistical model of diagnostic practice, and suggests sources for error, This model further injects noted types of human error and heuristics into the cognitive problem.

Reconciling this model of diagnosis is the job of the physician oriented towards practicing the art of medicine. In brief, akin to reconciling the mind-body problem and defining disease, is the reconciliation of the two facets of medicine whether they be described as: 1. clinical and the laboratory 2. linguistic and statistical 3. describable and ontological 4. psychosocial and biomedical

*Canguilhem* contrasts the ontological and the describable - or the statistical and the linguistic and suggests a model of disease that integrates both theories. This model seeks to diagnose pathological perturbation, and render treatment to return the patient to a pathological norm. With the aim and need to incorporate the context and nature of disease both, we can see how statistical and linguistic mechanisms serve to build a backbone of theory and conceptual understanding about the practice of medicine. Furthermore, given the infomedical model of disease and the current paradigm of science, information theoretic concepts and in turn the field of informatics, serve as a potential starting point for future advancements and improvements of care.

- Ledley, R. S., & Lusted, L. B. (1959). Reasoning Foundations of Medical Diagnosis. Science, 130(3366).
- Feinstein, A. R. (1973). An analysis of diagnostic reasoning. I. The domains and disorders of clinical macrobiology. The Yale Journal of Biology and Medicine, 46(3), 212.
- Feinstein, A. R. (1973). An analysis of diagnostic reasoning. II. The strategy of intermediate decisions. The Yale Journal of Biology and Medicine, 46(4), 264.
- Feinstein, A. R. (1974). An analysis of diagnostic reasoning. III. The construction of clinical algorithms. The Yale Journal of Biology and Medicine, 47(1), 5.
- Feinstein, A. R. (1994). Clinical Judgment Revisited: The Distraction of Quantitative Models. Annals of Internal Medicine, 120(9), 799–805.
- Croskerry, P. (2009). A universal model of diagnostic reasoning. Academic Medicine, 84(8), 1022–1028.
- Norusis, M. J., & Jacquez, J. A. (1975a). Diagnosis. I. Symptom nonindependence in mathematical models for diagnosis. Computers and Biomedical Research, 8(2), 156-172.
- Norusis, M. J., & Jacquez, J. A. (1975b). Diagnosis. II. Diagnostic models based on attribute clusters: A proposal and comparisons. Computers and Biomedical Research, 8(2), 173-188.
- Szolovits, P., & Pauker, S. G. (1976). Research on a medical consultation system for taking the present illness. In Proceedings of the Third Illinois Conference on Medical Information Systems, 299 (Vol. 320).
- Szolovits, P., & Pauker, S. G. (1978). Categorical and probabilistic reasoning in medical diagnosis. Artificial Intelligence, 11(1), 115–144.
- Canguilhem, G., with Fawcett C. R. & Cohen R. S. (1978). On the Normal and the Pathological. D Reidel Publishing Company.

### The Origins of Medical Informatics, Definition of Biomedical Informatics

Questions remain as to the nature of information theory, informatics and the concepts that have only lightly been touched on in this module. To whit, this section serves to describe the history of the field of medical informatics, and the Regenstrief Institute’s role in that history.

*Shortliffe and Blois* in their seminal treatise, describe the term **medical informatics**, as having originated in Europe. *Collen* furthers this idea, with his commentary in the Western Journal of Medicine, as do other fellows of the American College of Medical Informatics in the issue. This term applies to the application of the field of computer science to medical topics, as well as medical statistics, records keeping, and information science and study. In the United States, this definition was accepted, however, the term’s utility as naught but an awkward neologism was, and continues to be debated. As such, the general domain of biomedical informatics was fashioned, intending to encompass bioinformatics, imaging informatics, clinical informatics and public health informatics. With the expansion of the field and new applications envisioned, the term health informatics has been proposed as a more *meaningful* term that encompasses all facets of health care, including allied health, public health, and consumer healthcare.

The distinctions between fields and disciplines in the academy and industry are an artefact of the university culture and industrial culture in the country to which one examines them, and as such, the nomenclature of the United States is currently vacillating between biomedical informatics and health informatics, typically with medical school supported units favouring the former, while units supported by other institutions and only tangentially related to nursing/allied health favouring the latter. The American Medical Informatics Association (AMIA), defines biomedical informatics (with four corollaries) as:

the interdisciplinary field that studies and pursues the effective uses of biomedical data, information, and knowledge for scientific inquiry, problem solving, and decision making, motivated by efforts to improve human health.

1. develops, studies and applies theories, methods and processes for the generation, storage, retrieval, use, and sharing of biomedical data, information, and knowledge.
2. builds on computing, communication and information sciences and technologies and their application in biomedicine.
3. investigates and supports reasoning, modeling, simulation, experimentation and translation across the spectrum from molecules to populations, dealing with a variety of biological systems, bridging basic and clinical research and practice, and the healthcare enterprise.
4. recognizing that people are the ultimate users of biomedical information, draws upon the social and behavioral sciences to inform the design and evaluation of technical solutions and the evolution of complex economic, ethical, social, educational, and organizational systems.

*Dick, Steen, Detmer et al* authored a report in 1997 argued for the adoption of computer-based patient records, in support of easily collecting information for clinical trials, as well as improving care. As discussed by *Weed* in the luminary series, the role of the medical record and the advantages of establishing a closed loop system using a computer to store and represent information is valuable. *Friedman* has taken this further in current policy literature, with the concept of the learning health care system, as well as his fundamental theory of biomedical informatics. Biomedical informatics, and indeed, the entirety of the American philosophy towards the discipline (as is alluded to by *Shortliffe and Blois*) has been dominated by the practical, rather than the theoretical. Within the current health information technology landscape, the growth in*health informatics* and like programs reflect that focus. Therefore, outside of the oldest institutions, the definition of medical informatics and its practice are dominated by pseudo-engineering processes and methods, primarily shaped by management science and the business school. This is exemplified in the discussion of biomedical engineering by *Shortliffe and Blois*, and their inaccuracy in the identification of the discipline, versus clinical engineering, let alone the practice of engineering. Clinical engineering, a discipline which has primarily applied its efforts to medical devices rather than information technology, has seen its recent capacity in the information technology and informatics domains shift with the publication of the joint National Academy of Engineering and Institute of Medicine report entitled *Building a Better Delivery System: A New Engineering/Health Care Partnership*, and the writings of *Zambuto and Grimes*. As one can surmise, the exact nature of biomedical informatics, the role of theory and application, and even the terminology of the discipline are currently debated and malleable topics. American medical informatics, and its current practice and the society to which it belongs, is moving at a pace constantly changed by market, academic and clinical forces.

- Shortliffe, E. H., & Blois, M. S. (2001). The computer meets medicine and biology: emergency of a discipline. Biomedical Informatics: Computer Applications in Healthcare and Biomedicine. New York: Springer-Verlag.
- Collen, M. F. (1986). Origins of medical informatics. Western Journal of Medicine, 145(6), 778.
- Dick, R. S., Steen, E. B., Detmer, D. E., & others. (1997). The Computer-Based Patient Record: An Essential Technology for Health Care (Revised.). National Academies Press.
- Friedman, C. P., Wong, A. K., & Blumenthal, D. (2010). Achieving a nationwide learning health system. Science translational medicine, 2(57), 1-3.
- Friedman, C., Rubin, J., Brown, J., Buntin, M., Corn, M., Etheredge, L., & Van Houweling, D. (2014). Toward a science of learning systems: a research agenda for the high-functioning Learning Health System. Journal of the American Medical Informatics Association 0() 1-6.
- Friedman, C. P. (2009). A “fundamental theorem” of biomedical informatics. Journal of the American Medical Informatics Association, 16(2), 169-170.
- Fanjiang, G., Grossman, J. H., Compton, W. D., & Reid, P. P. (Eds.). (2005). Building a Better Delivery System: A New Engineering/Health Care Partnership. National Academies Press.
- Zambuto, R., &amp; Grimes, S. (2010). The Growing Move Toward Clinical Systems Engineering. Biomedical Instrumentation & Technology, 44(5), 426-432.

### A Brief History of the Regenstrief Institute and the Clinical Informatics Specialty

The Regenstrief Institute was founded as a “laboratory for research, integrating research programs into actual patient care” in 1967. It currently stands as an affiliate research institute of the Indiana University School of Medicine. Since 1972, the institute as been the birthplace of the concept of the Health Information Exchange with the *Indiana Network for Patient Care*, laboratory and measurement coding with *Logical Observation Identifiers Names and Codes*, and the medical record system with the *Regenstrief Medical Record System* and *Gopher*. It serves as one of the birthplaces of clinical informatics in the former Wishard Memorial Hospital (now Eskenazi Health) in internal medicine and primary care. In 2000, Regenstrief’s researchers were instrumental in founding [ResNet](https://www.indianactsi.org/programs/researchnetworks/resnet), a university-affiliated clinician-researcher partnership to answer care-based decisions. It stands as a practice-based research network, serving the entire Tri-Institution [Clinical and Translational Sciences Institute](https://www.indianactsi.org) state-wide. This approach, championed by Sam Regenstrief himself, focused on the improvement of healthcare through practice and research. The legacy of Sam Regenstrief, as described in the titular book, was to use technological innovation to improve the quality and delivery of health care in the same way he used technological innovation to advance the Original Equipment Manufacturer (OEM) dishwasher industry.

The Regenstrief Foundation, which funds the institute has had its mission [described](https://www.purdue.edu/discoverypark/rche/about/who.php) as:

to provide support for innovative research directed toward improving the efficiency, quality, and accessibility of healthcare. We will emphasize research in informatics, epidemiology, economics, and interventions in healthcare delivery. We will support selected programs by providing sustained long-term support of institutional partners. In pursuing this mission, we are committed to building on Sam Regenstrief’s legacy by stressing creativity, adaptability, and results in both what we do and how we operate.

Other innovations, such as the *Child Health Improvement through Computer Automation* (CHICA) system, have been produced by Regenstrief investigators alongside its core operations within the living laboratory that is Eskenazi Hospital. This system offers patient and provider tailored clinical decision support for the pediatric population within the local academic pediatric practices, as well as the capacity to easily support clinical reasoning and care-focused trials through PResNet (Pediatric Research Network), which serves as a focal point or research in the department of pediatrics, much like ResNet does for the adult and TeenNet for the adolescent population. Regenstrief serves as the home for major leaps forward in clinical-facing technology, which provide it the opportunity to advance point-of-care medical delivery, but also to perform research on the medical information collected during the course of care: both for specific research purposes via technology, and also via the normative practice of electronic clinical documentation. To perform these tasks, the institute operates within the academic discipline of informatics, but also that of health services research. Health services research is a discipline [defined](http://www.nlm.nih.gov/nichsr/ihcm/01whatis/whatis07.html) by the National Information Center on Health Services Research and Health Care Technology (NICHSR) as:

[investigating] three major aspects of health care: access to care, the quality of the care, and its cost. Health services researchers attempt to evaluate the effects and outcomes of the health care “system” on people’s health.

Health services research offers a mechanism by which researchers can investigate facets of clinical operations, reasoning and practice; and in the case of the Regenstrief Institute, all supported by technological innovation. Clinical informatics is described by the Accreditation Council for Graduate Medical Education in its program requirements (taken from *Gardner et al*) as:

[the] subspecialty of all medical specialties that transforms health care by analyzing, designing, implementing, and evaluating information and communication systems to improve patient care, enhance access to care, advance individual and population health outcomes, and strengthen the clinician-patient relationship

The [American Board of Preventive Medicine](http://www.theabpm.org/examinfo-ci.cfm) administrates the clinical informatics examination for board certification. The core content for the sub speciality was described first in 2009 by *Gardner et al*, and has been further refined in its report on program requirements. Regenstrief has been instrumental in developing this core content, and along with Stanford University and Oregon Health and Sciences University, is first in the nation for adapting its previously existing fellowship program (since approximately 1975) to support the board-certification process and credentialing of physicians.

- Ford, Wendy, Joanne Fox, and Julie Sturgeon. Regenstrief: Legacy of the Dishwasher King. Regenstrief Foundation, 1999.
- Gardner, R. M., Overhage, J. M., Steen, E. B., Munger, B. S., Holmes, J. H., Williamson, J. J., … & AMIA Board of Directors. (2009). Core content for the subspecialty of clinical informatics. Journal of the American Medical Informatics Association, 16(2), 153-157.
- Accreditation Council for Graduate Medical Education, “ACGME Program Requirements for Graduate Medical Education in Clinical Informatics,” ACGME, Chicago, IL, 2016
