## Supplemental File and Appendix for "Designing a Clerkship Curriculum for Medical Students in Clinical and Medical Informatics in the Electronic Medical Record Era": HIPAA Training - Pages Concatenated.docx

### The Culture of Risk

The culture of risk that pervades the health information technology (HIT) field primarily has to do with legal risk and regulation concerning the passage of two laws:

1. HITECH
2. HIPAA

These laws concern the use, transmission and general dealings with electronic personal health information (EPHI). EPHI is personally identifiable health information in electronic form. Whether this information is protected or not leads to the application of the HIPAA privacy or security rules.

- The HIPAA privacy rule, titled *Privacy of Individually Identifiable Health Information*, covers protected health information in its use in any form.
- The HIPAA security rule, *Security Standards for the Protection of Electronic Protected Health Information*, covers electronically protected health information.

Examples of protected health information from [USC Tile 45 Subtitle A Subchapter C Part 164 Subpart E §164.501](http://www.ecfr.gov/cgi-bin/text-idx?SID=efc1ee374e58623f65fcb55afe4d2948&mc=true&node=sp45.1.164.e&rgn=div6#se45.1.164_1501) include:

- Name and address;
- Date of birth;
- Social security number;
- Payment history;
- Account number; and
- Name and address of the health care provider and/or health plan.

#### Definitions

- *Health Information* described in [Public Law 104 - 191](http://www.gpo.gov/fdsys/pkg/PLAW-104publ191/content-detail.html) as:
- The term ‘health information’ means any information, whether oral or recorded in any form or medium, that—
- ‘‘(A) is created or received by a health care provider, health plan, public health authority, employer, life insurer, school or university, or health care clearinghouse; and
- ‘‘(B) relates to the past, present, or future physical or mental health or condition of an individual, the provision of health care to an individual, or the past, present, or future payment for the provision of health care to an individual.
- *Individually identifiable health information* is described in [Public Law 104 - 191](http://www.gpo.gov/fdsys/pkg/PLAW-104publ191/content-detail.html) as:
- The term ‘individually identifiable health information’ means any information, including demographic information collected from an individual, that—
- ‘‘(A) is created or received by a health care provider, health plan, employer, or health care clearinghouse; and
- ‘‘(B) relates to the past, present, or future physical or mental health or condition of an individual, the provision of health care to an individual, or the past, present, or future payment for the provision of health care to an individual, and—
- ‘‘(i) identifies the individual; or
- ‘‘(ii) with respect to which there is a reasonable basis to believe that the information can be used to identify the individual.

### The HITECH Act

The Health Information Technology for Economic and Clinical Health (HITECH) act, enacted as part of the American Recovery and Reinvestment Act (ARRA) in 2009 has a subtitle: [Subtitle D](http://www.gpo.gov/fdsys/search/pagedetails.action?collectionCode=USCODE&searchPath=Title+42%2FChapter+156%2FSUBCHAPTER+III&oldPath=Title+42%2FChapter+156%2FSUBCHAPTER+III&isCollapsed=false&selectedYearFrom=2013&ycord=6400&browsePath=Title+42%2FChapter+156%2FSubchapter+III%2FSec.+17921&granuleId=USCODE-2013-title42-chap156-subchapIII-sec17921&packageId=USCODE-2013-title42&collapse=true&fromBrowse=true), which directly addresses privacy and security concerns relating to electronic health records. This subtitle defines the terms:

1. *breach*:

The term “breach” means the unauthorized acquisition, access, use, or disclosure of protected health information which compromises the security or privacy of such information, except where an unauthorized person to whom such information is disclosed would not reasonably have been able to retain such information.

2. *exceptions*:

The term “breach” does not include -

1. any unintentional acquisition, access, or use of protected health information by an employee or individual acting under the authority of a covered entity or business associate if —
   1. such acquisition, access, or use was made in good faith and within the course and scope of the employment or other professional relationship of such employee or individual, respectively, with the covered entity or business associate; and
   2. such information is not further acquired, accessed, used, or disclosed by any person; or
2. any inadvertent disclosure from an individual who is otherwise authorized to access protected health information at a facility operated by a covered entity or business associate to another similarly situated individual at 1 same facility; and
3. any such information received as a result of such disclosure is not further acquired, accessed, used, or disclosed without authorization by any person.

### The MACR Act of 2015

[H.R.2, the Medicare Access and CHIP Reauthorization Act of the 114th Congress](https://www.congress.gov/bill/114th-congress/house-bill/2/text)was issued as a means

To amend title XVIII of the Social Security Act to repeal the Medicare sustainable growth rate and strengthen Medicare access by improving physician payments and making other improvements, to reauthorize the Children's Health Insurance Program, and for other purposes.

Section 106 "REDUCING ADMINISTRATIVE BURDEN AND OTHER PROVISIONS" Subsection (b) re-energizes the commitment of the government to EMR interoperability.

Specifically, it states the objective as

As a consequence of a significant Federal investment in the implementation of health information technology through the Medicare and Medicaid EHR incentive programs, Congress declares it a national objective to achieve widespread exchange of health information through interoperable certified EHR technology nationwide by December 31, 2018.

This acknowledges the circumstances surrounding HIT in 2015/2016, and suggests that HIPA and other acts will only become more important with time.

Furthermore, in a [press release](http://www.ama-assn.org/ama/pub/news/news/2016/2016-06-03-interoperability-measurements-ehr.page) on June 3rd 2016, the American Medical Association acknowledged that the landscape has indeed changed, and along with 36 medical specialty associations has put forward a call to "make the promise of the EMR a reality".

### The HIPA Act

The Health Insurance Portability and Accountability Act (HIPAA) sets out a security standard, and provides for the culture of risk currently present in the care environment.

- The Department of Health and Human Services (DHHS) maintains the combined text of the administrative simplifications afforded for by HIPAA: [Regulation Combined Text](http://www.hhs.gov/ocr/privacy/hipaa/administrative/combined/index.html).
- Furthermore the National Institute of Standards and Technology has issued a [HIPAA security rule toolkit](http://scap.nist.gov/hipaa/), which serves as a resource to support an organization’s risk assessment process.
- The Office of the National Coordinator for Health Information Technology (ONC) alongside the HHS Office for Civil Rights (OCR) and Office of the General Counsel (OGC) has developed a [security risk assessment tool](http://www.healthit.gov/providers-professionals/security-risk-assessment-tool)

[HIPAA Security and Privacy](http://www.ecfr.gov/cgi-bin/text-idx?tpl=/ecfrbrowse/Title45/45cfr164_main_02.tpl) is covered by USC Title 45, Subtitle A, Subchapter C, Part 164.

- The [HIPAA Security Rule](http://www.ecfr.gov/cgi-bin/text-idx?SID=efc1ee374e58623f65fcb55afe4d2948&mc=true&node=sp45.1.164.c&rgn=div6) §164.304 describes three key terms:
  1. *Confidentiality*
  - the property that data or information is not made available or disclosed to unauthorized persons or processes
  2. *Integrity*
  - the property that data or information have not been altered or destroyed in an unauthorized manner.
  3. *Availability*
  - the property that data or information is accessible and useable upon demand by an authorized person
- The [HIPAA Privacy Rule](http://www.ecfr.gov/cgi-bin/text-idx?SID=efc1ee374e58623f65fcb55afe4d2948&mc=true&node=sp45.1.164.e&rgn=div6) §164.501 describes general and organizational rules for uses and disclosures of protected health information in §164.502,164.504 alongside rules related to carrying out:
  1. Treatment operations
  2. Payment operations
  3. Health care operations

These fall into several categories:

1. Authorization is required
2. An opportunity for the individual to agree or to object is required
3. An opportunity for the individual to agree or to object is not required

These categories assist in defining the access rights of individuals, accounting requirements, administrative requirements and subsequently translate to security requirements in terms of administrative, physical and technical safeguards. These safeguards and the implementation of the Security Rule have particular guidelines.

The [NIST Computer Security Division](http://csrc.nist.gov/) has published a resource guide for implementing HIPAA security rule (§164.306(a)), supporting each covered entity in their requirement to:

1. Ensure the confidentiality, integrity, and availability of EPHI that it creates, receives, maintains, or transmits
2. Protect against any reasonably anticipated threats and hazards to the security or integrity of EPHI; and
3. Protect against reasonably anticipated uses or disclosures of such information that are not permitted by the Privacy Rule.

### Data at Rest vs Data in Motion

The concepts of data at rest and data in motion, offers two distinct categories by which information can be contained from a technological standpoint. As such, IT departments often refer to these concepts in there documentation, policies and conversations.

- *At Rest*:
  - Data kept in a physical storage device, not being transmitted
- *In Motion*:
  - Data moving through a transmission channel (computer network, ADSL line…)

### The Department of Health and Human Services Office of Civil Rights

This office serves as a one stop shop for both [federal civil rights claims](http://www.hhs.gov/ocr/civilrights/index.html) and [health information privacy](http://www.hhs.gov/ocr/privacy/index.html). With regards to health information privacy, this office enforces:

- [HIPAA Privacy Rule](http://www.ecfr.gov/cgi-bin/retrieveECFR?gp=&SID=45a928fb311efaf37ee9c087d45dd0d9&mc=true&n=pt45.1.164&r=PART&ty=HTML#sp45.1.164.e)
- [HIPAA Security Rule](http://www.ecfr.gov/cgi-bin/retrieveECFR?gp=&SID=45a928fb311efaf37ee9c087d45dd0d9&mc=true&n=pt45.1.164&r=PART&ty=HTML#sp45.1.164.c)
- [HIPAA Breach Rule](http://www.ecfr.gov/cgi-bin/retrieveECFR?gp=&SID=45a928fb311efaf37ee9c087d45dd0d9&mc=true&n=pt45.1.164&r=PART&ty=HTML#sp45.1.164.d)
- Patient Safety Rule, a component of the [Patient Safety and Quality Improvement Act of 2005](http://www.gpo.gov/fdsys/pkg/PLAW-109publ41/content-detail.html)

### Breaches Affecting 500 or More Individuals

#### Breaches Affecting 500 or More Individuals

The HIPAA Breach Rule, formally entitled *Notification in the Case of Breach of Unsecured Protected Health Information*, requires the formal notification of several entities for an event of greater than 500 persons:

1. Individuals affected
2. Media serving the jurisdiction
3. Secretary of Health and Human Services

Furthermore, due to section 13402(e)(4) of HITECH, the Secretary of Health and Human Services must post a list of said breaches. This information is stored on a website maintained by the [Department of Health and Human Services](http://ocrportal.hhs.gov/ocr/breach/breach_report.jsf) The following code performs statistics on the aggregate data file obtainable at the aforementioned URL.

##### R Code for Brief Statistics on the [Breaches Affecting 500 or More Individuals File](http://ocrportal.hhs.gov/ocr/breach/breach_report.jsf)

- Download CSV file from website (breach_report.csv)

```
breach_report <- read.csv('breach_report.csv',header=T)
#Frequency of Breach Reports by State
table(breach_report$State)
##
##      AK  AL  AR  AZ  CA  CO  CT  DC  DE  FL  GA  HI  IA  ID  IL  IN  KS
##   8   5  17   7  26 141  21  18   7   1  77  46   1   7   3  65  44   9
##  KY  LA  MA  MD  ME  MI  MN  MO  MS  MT  NC  ND  NE  NH  NJ  NM  NV  NY
##  27  11  37  21   1  25  30  26   6   7  35   3   6   4  18  11  11  80
##  OH  OK  OR  PA  PR  RI  SC  SD  TN  TX  UT  VA  VT  WA  WI  WV  WY
##  37   9  19  49  28   7  13   2  34 110  11  23   1  29  11   5   4
plot(breach_report$State,las=2)
```

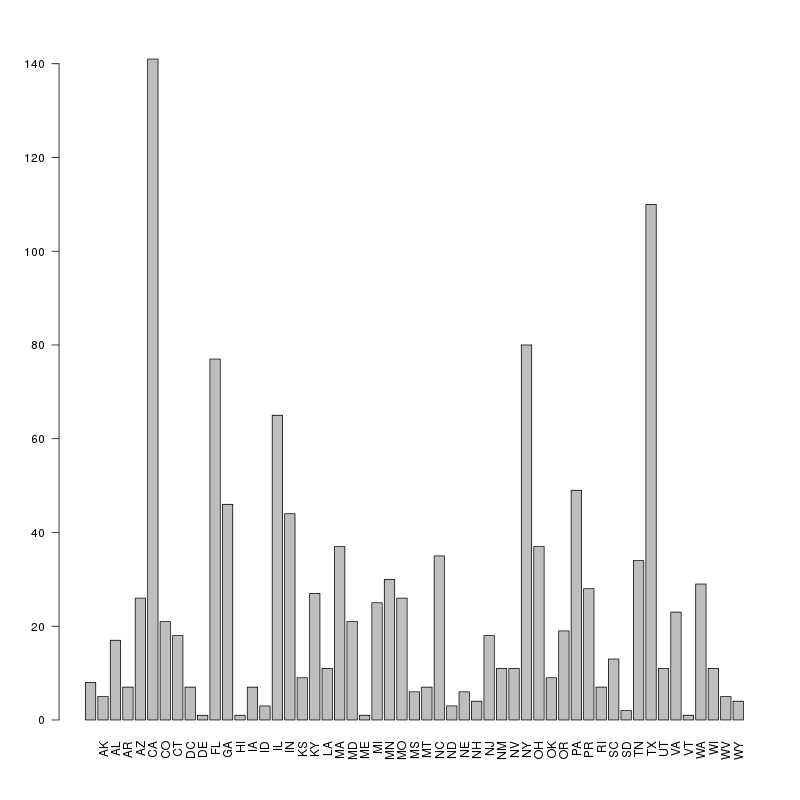

```
#Frequency of Breach Reports by Covered Entity Type
table(breach_report$Covered.Entity.Type)
##
##                                  Business Associate
##                         2                       275
## Healthcare Clearing House       Healthcare Provider
##                         4                       836
##               Health Plan
##                       137
#set margins
par(mar=c(13,5,1,1))
plot(breach_report$Covered.Entity.Type,las=2)
```

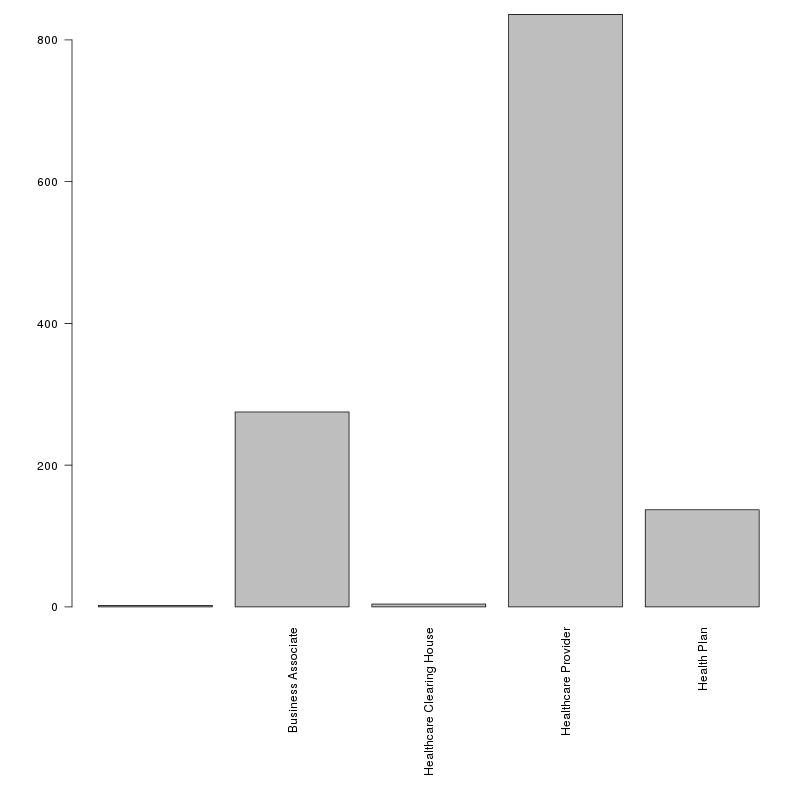

```
#Summary Statistics for Number of Individuals Affected per Breach
summary(breach_report$Individuals.Affected)
##     Min.  1st Qu.   Median     Mean  3rd Qu.     Max.
##      500      985     2394   107600     7458 78800000
```

### IU Requirements

Institutionally, as a partner with Indiana University Health, and as the School of Medicine, there are several mechanisms by which HIPAA enforcement and requirements govern the actions of students, staff and faculty.

- Compliance with HIPAA at IU is coordinated through the Office of the Vice President and General Counsel
- IU is a covered entity with hybrid status
  - Single legal entity, with components both covered and non-covered
  - Covered entities are those that access PHI (e.g. healthcare provider, health plan), provide services to covered components (and receives PHI to perform tasks) or uses PHI for education
  - UITS Advanced Biomedical IT Core provides services for researchers who need help dealing with PHI

#### Resources

- Knowledge Base: [What is HIPAA](https://kb.iu.edu/d/ayyy)
- Knowledge Base: [At IU, how do I get help securely processing, storing, and sharing ePHI or HIPAA data?](https://kb.iu.edu/d/ayzb)
- Office of the Vice President and General Counsel: [HIPAA Compliance Education](http://www.iub.edu/%7Evpgc/compliance/hipaa-privacy-and-security/hipaa-compliance-education.shtml)
- HIPAA Module via [E-Training](https://one.iu.edu/task/iu/e-training)

### Regenstrief as a Covered Entity

Due to our data agreements, we serve as a covered entity

- Primarily due to hosting the Indiana Network for Patient Care - Regenstrief clone for research and service purposes

As with IU, Regenstrief is responsible for coverage related to:

1. Privacy requirements
2. Security Requirements
3. Breach requirements

Given Regenstrief’s position solely as a covered entity, without breaks relating to non-health related topics, the organization’s operationalization of these requirements are far stricter and more controlled than IU’s.

### Legal Risk as a Covered Entity

As a result of these requirements, particularly the risk of a data breach, legal paranoia sets in. Data use agreements and business associate agreements allow for formalizing privacy and security requirements under contract law. Tort law is a primary mechanism to govern breaches of privacy, yet the details of prosecution or even liability have not been sorted out. Other elements of digital presence that intersect with the medical sphere (such as social media) may play a part in increasing liability. Legal literature on the topic supports controlling interaction between covered entities and the outside world, and has led to restrictive social media policies in hospitals, strenuous data encryption and use requirements, and restrictions on research activities and questions due to prior agreements or data use policies.

##### Readings

- Brill, Jack. "Giving HIPAA Enforcement Room to Grow: Why There Should Not (Yet) Be a Private Cause of Action." Notre Dame L. Rev. 83 (2007): 2105. (A)
- Collins, Joshua DW. “Toothless HIPAA: Searching for a private right of action to remedy privacy rule violations.” Vand. L. Rev. 60 (2007): 199. (A)
- Pasternack, Eric S. "HIPAA in the Age of Electronic Health Records." Rutgers LJ 41 (2009): 817.
- Terry, Nicolas. "Physicians and Patients Who "Friend" or "Tweet": Constructing a Legal Framework for Social Networking in a Highly Regulated Domain." Indiana Law Review 43, no. 285 (2010). (A)

### Assignment: CITI Training

The [Office of Research Administration](http://researchcompliance.iu.edu/eo/eo_citi.html) provides the education portal to the [Collaborative Institutional Training Initiative](http://citi.iu.edu/?submit=Yes)

Provide us the completion report from this initiative for the following courses from Indiana University - Indianapolis:

1. Good Clinical Practice

2. Biomedical Researcher

3. Biomedical Responsible Conduct of Research

4. Social/Behavioral Reseachers

5. Social and Behavioral Responsible Conduct of Research

### Assignment: (Certificate of Completion) HIPAA Privacy & Security - IN Medical Students

Please provide us with a screenshot of the certificate of completion.

Log in to E-Training from one.iu.edu <[https://one.iu.edu/task/iu/e-training](http://one.iu.edu/task/iu/e-training)>
