## Supplemental File and Appendix for "Designing a Clerkship Curriculum for Medical Students in Clinical and Medical Informatics in the Electronic Medical Record Era": Luminary Paper Series - Pages Concatenated.docx

### Larry Weed

1. Weed, L. L. (1964). Medical Records, Patient Care and Medical Education. Irish Journal of Medical Science, 39(6), 271–282. (A)
2. Weed, L. L. (1968). Medical records that guide and teach. New England Journal of Medicine, 278(12), 593–600. (A)
3. Weed, L. L., & Weed, L. (2011). Medicine in Denial. CreateSpace. (B)

#### The SOAP Note

1. Moser, E. M., Huang, G. C., Packer, C. D., Glod, S., Smith, C. D., Alguire, P. C., & Fazio, S. B. (2015). SOAP-V: Introducing a method to empower medical students to be change agents in bending the cost curve. Journal of Hospital Medicine.
2. Lin CT, McKenzie M, Pell J, Caplan L. Health care provider satisfaction with a new electronic progress note format: SOAP vs APSO format. JAMA internal medicine. 2013 Jan 28;173(2):160-2.
3. Shoolin J, Ozeran L, Hamann C, Bria II W. Association of Medical Directors of Information Systems consensus on inpatient electronic health record documentation. Applied clinical informatics. 2013;4(2):293.
4. Clarke MA, Steege LM, Moore JL, Koopman RJ, Belden JL, Kim MS. Determining primary care physician information needs to inform ambulatory visit note display. Applied clinical informatics. 2013 Dec;5(1):169-90.
5. Hahn JS, Bernstein JA, McKenzie RB, King BJ, Longhurst CA. Rapid implementation of inpatient electronic physician documentation at an academic hospital. Appl Clin Inform. 2012 Jan 1;3(2):175-85.
6. Brown PJ, Marquard JL, Amster B, Romoser M, Friderici J, Goff S, Fisher D. What do physicians read (and ignore) in electronic progress notes?. Applied clinical informatics. 2014;5(2):430.

Weed’s creation of the SOAP note has been suggested as potentially needing modification in the current age of the EMR. To which, flipping the Assessment and Plan to before the Subject and Objective has been suggested (APSO), as well as adding a Value component (SOAP-V). Weed has commented on the former in an issue of Healthcare IT News [Versel, N. Physicians rethinking the progress note. Healthcare IT News. 2014 May 8](http://www.healthcareitnews.com/news/rethinking-progress-note), saying on the topic that:

Never put your opinion before the facts. The supposed advantage of the APSO alternative – that it begins with the physician’s assessment rather than data – is actually a failing. This sequence tends to make the note provider-centered rather than patient-centered, and judgment-based rather than evidence-based. In contrast, beginning the progress note with data disciplines the provider’s assessment. The provider must think in terms of specific data, specific problems on the problem list to which the data relate, and the interrelationship of each problem to the other problems on the list. Moreover, it’s important to begin the progress note with subjective (symptomatic) data from the patient rather than so-called ‘objective’ data.

Commensurate with the articles commenting on the value of APSO, Epic has built out the ability to provide a collapsed view of the SOAP components of the note as demonstrated by the following two screenshots from both Epic documentation and Ezkenazi’s production system.

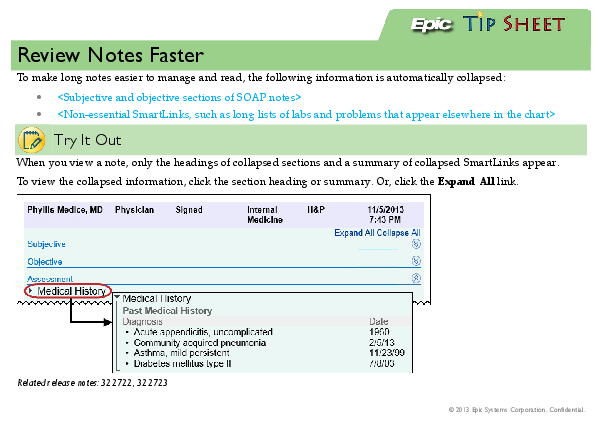

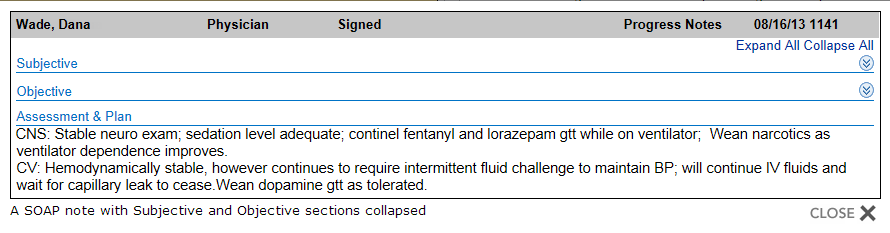

At the same time as the SOAP note’s birth, *Engel and Morgan*, the former of biopsychosocial model fame, discuss their thoughts on the recording of the note and its contents in an excerpt of Chapter 5 and Appendix A from their seminal text *The Clinical Approach to the Patient*. This excerpt comes around the same time as Weed developed the concepts of the SOAP note and its implementation, and comes with its own history from the University of Rochester’s medical education research from 1944-1966.

1. Morgan WL, Engel GL. The clinical approach to the patient. WB Saunders Company; 1969
2. Engel GL. Medical education and the psychosomatic approach: A report on the Rochester experience 1946–1966. Journal of psychosomatic research. 1967 Jun 30;11(1):77-85.

### Clement McDonald, Marc Overhage and Bill Tierney

1. *McDonald, Clement J., J. Marc Overhage, Paul R. Dexter, Lonnie Blevins, Jim Meeks-Johnson, Jeffrey G. Suico, Mark C. Tucker, and Gunther Schadow. "Canopy computing: using the Web in clinical practice." Jama 280, no. 15 (1998): 1325-1329.
2. *McDonald, C. J. (1976). Protocol-Based Computer Reminders, the Quality of Care and the Non-Perfectibility of Man. New England Journal of Medicine, 295(24), 1351–1355. (A)
3. *Tierney, W. M., Overhage, J. M., & J., M. C. (1995). Toward electronic medical records that improve care. Annals of Internal Medicine, 9, 725. (A)
4. McDonald CJ, Huff SM, Suico JG, Hill G, Leavelle D, Aller R, Forrey A, Mercer K, DeMoor G, Hook J, Williams W, Case J, Maloney P. LOINC, a universal standard for identifying laboratory observations: a 5-year update. Clin Chem. 2003 Apr;49(4):624-33.
5. Huff, S. M., Rocha, R. A., McDonald, C. J., De Moor, G. J., Fiers, T., Bidgood, W. D., … & Baenziger, J. (1998). Development of the logical observation identifier names and codes (LOINC) vocabulary. Journal of the American Medical Informatics Association, 5(3), 276-292.
6. Vreeman DJ, McDonald CJ. Automated mapping of local radiology terms to LOINC. AMIA Annu Symp Proc. 2005:769-73.
7. Vreeman DJ, Finnell JT, Overhage JM. A rationale for parsimonious laboratory term mapping by frequency. AMIA Annu Symp Proc. 2007 Oct 11:771-5.
8. McDonald, C., Bhargava, B., & Jeris, D. (1975, May). A clinical information system (CIS) for ambulatory care. In Proceedings of the May 19-22, 1975, national computer conference and exposition (pp. 749-756). ACM.
9. *Tierney, W. M., Miller, M. E., Overhage, J. M., & McDonald, C. J. (1993). Physician inpatient order writing on microcomputer workstations: effects on resource utilization. Jama, 269(3), 379-383.
10. *McDonald, C. J. (1996). Medical heuristics: the silent adjudicators of clinical practice. Annals of Internal Medicine, 124(1:1), 56-62.
11. *McDonald, C. J. (1976). Use of a computer to detect and respond to clinical events: its effect on clinician behavior. Annals of Internal Medicine, 84(2), 162-167

* = Required reading

### Edward Shortliffe

1. Shortliffe, E. H., & Buchanan, B. G. (1975). A Model of Inexact Reasoning in Medicine. Mathematical Biosciences, 23, 351–379. (A)
2. Clancey, W. J., & Shortliffe, E. H. (1984). Readings in medical artificial intelligence: the first decade. Addison-Wesley Longman Publishing Co., Inc. (B)
3. Shortliffe, E. H. (1976). MYCIN: Computer-based medical consultations. Elsevier, New York. (B)
4. Buchanan, B. G., & Shortliffe, E. H. (1984). Rule Based Expert Systems: The Mycin Experiments of the Stanford Heuristic Programming Project. Addison-Wesley Longman Publishing Co., Inc. (B)

### Octo Barnett

### Naomi Sager and Carol Friedman

1. Friedman, C., Kra, P., & Rzhetsky, A. (2002). Two biomedical sublanguages: a description based on the theories of Zellig Harris. Journal of Biomedical Informatics, 35(4), 222–235. (A)
2. * Friedman, C., & Johnson, S. B. (2013). Natural language and text processing in biomedicine. In Biomedical Informatics: Computer Applications in Health Care and Biomedicine. Springer New York. (B)
3. Sager, N., Friedman, C., & Lyman, M. S. (1987). Medical Language Processing: Computer Management of Narrative Data. Addison-Wesley. (B)
4. Sager, N. (1979). Natural Language Information Formatting: The Automatic Conversion of Texts to a Structured Data Bases. Advances in Computers, 17, 89. (A)
5. Sager, N., Lyman, M., Bucknall, C., Nhan, N., & Tick, L. J. (1994). Natural Language Processing and the Representation of Clinical Data. Journal of the American Medical Informatics Association, 1(2), 142-160. (A)

### Titus Schleyer

1. Acharya, A., Mital, D. P., & Schleyer, T. K. (2009). Electronic dental record information model. International Journal of Medical Engineering and Informatics, 1(4), 418-434. (A)
2. Schleyer, T. (2007). Dentists, patients and medications. The Journal of the American Dental Association, 138(2), 148. (A)
3. Schleyer, T. K., Thyvalikakath, T. P., Spallek, H., Torres-Urquidy, M. H., Hernandez, P., & Yuhaniak, J. (2006). Clinical computing in general dentistry. Journal of the American Medical Informatics Association, 13(3), 344-352. (A)
4. Schleyer, T. K., Corby, P., & Gregg, A. L. (2003). A preliminary analysis of the dental informatics literature. Advances in dental research, 17(1), 20-24. (A)
5. Schleyer, T. K. (2003). Dental informatics: an emerging biomedical informatics discipline. Advances in Dental Research, 17(1), 4-8. (A)
6. Schleyer, T., & Spallek, H. (2001). Dental informatics: a cornerstone of dental practice. The Journal of the American Dental Association, 132(5), 605-613. (A)
7. Schleyer, T. K., Thyvalikakath, T. P., Spallek, H., Dziabiak, M. P., & Johnson, L. A. (2012). From information technology to informatics: the information revolution in dental education. Journal of dental education, 76(1), 142-153. (A)
8. Eisner, John. et al The computer-based oral health record: a new foundation for oral health information systems. University at Buffalo, School of Dental Medicine, 1993.
9. Schleyer, T. K., (2003). Dental Informatics &amp; Dental Research: Making the Connection, Proceedings of the Dental Informatics &amp; Dental Research: Making the Connection Conference, Bethesda, MD.

### Richard N Shiffman and Stephen M Downs

1. * Shiffman, R. N. (2009). Clinical practice guidelines: Supporting decisions, optimizing care. In Pediatric Informatics (pp. 149-159). Springer New York.
2. Hoeksema, L. J., Bazzy-Asaad, A., Lomotan, E. A., Edmonds, D. E., Ramírez-Garnica, G., Shiffman, R. N., & Horwitz, L. I. (2011). Accuracy of a computerized clinical decision-support system for asthma assessment and management. Journal of the American Medical Informatics Association, 18(3), 243-250.
3. Lomotan, E. A., Hoeksema, L. J., Edmonds, D. E., Ramírez-Garnica, G., Shiffman, R. N., & Horwitz, L. I. (2012). Evaluating the use of a computerized clinical decision support system for asthma by pediatric pulmonologists. International journal of medical informatics, 81(3), 157-165.
4. * Silverstone, D. E., Paek, H. M., Kogan, Y., Essaihi, A., & Shiffman, R. N. (2005). The Incorporation of Clinical Practice Guidelines for Glaucoma into an Ophthalmology Electronic Medical Record. In AMIA Annual Symposium Proceedings (Vol. 2005, p. 1115). American Medical Informatics Association.
5. Rosenfeld, R. M., Shiffman, R. N., & Robertson, P. (2013). Clinical Practice Guideline Development Manual, A Quality-Driven Approach for Translating Evidence into Action. Otolaryngology–Head and Neck Surgery, 148(1 suppl), S1-S55.
6. Lustig, J., Gotlieb, E. M., Deutsch, L., Gerstle, R., Lieberthal, A., Shiffman, R., … & Stern, M. (2001). Special requirements for electronic medical record systems in pediatrics. Pediatrics, 108(2), 513-515.
7. Benin, A. L., Vitkauskas, G., Thornquist, E., Shiffman, R. N., Concato, J., Krumholz, H. M., & Shapiro, E. D. (2003). Improving diagnostic testing and reducing overuse of antibiotics for children with pharyngitis: a useful role for the electronic medical record. The Pediatric infectious disease journal, 22(12), 1043-1047.
8. * Shiffman, R. N., Michel, G., Essaihi, A., & Marcy, T. W. (2004). Using a guideline-centered approach for the design of a clinical decision support system to promote smoking cessation. Studies in health technology and informatics, 152-156.
9. Anand, V., Biondich, P. G., Liu, G., Rosenman, M., & Downs, S. M. (2004). Child health improvement through computer automation: the CHICA system. Stud Health Technol Inform, 107(Pt 1), 187-191.
10. Biondich, P. G., Anand, V., Downs, S. M., & McDonald, C. J. (2003). Using adaptive turnaround documents to electronically acquire structured data in clinical settings. In AMIA Annual Symposium Proceedings (Vol. 2003, p. 86). American Medical Informatics Association.
11. * Downs, S. M., Biondich, P. G., Anand, V., Zore, M., & Carroll, A. E. (2006). Using Arden Syntax and adaptive turnaround documents to evaluate clinical guidelines. In AMIA Annual Symposium Proceedings (Vol. 2006, p. 214). American Medical Informatics Association.
12. Carroll, A. E., Biondich, P. G., Anand, V., Dugan, T. M., Sheley, M. E., Xu, S. Z., & Downs, S. M. (2011). Targeted screening for pediatric conditions with the CHICA system. Journal of the American Medical Informatics Association, 18(4), 485-490.
13. Downs, S. M., & Wallace, M. Y. (2000). Mining association rules from a pediatric primary care decision support system. In Proceedings of the AMIA Symposium (p. 200). American Medical Informatics Association.
14. * Downs, S. M., Zhu, V., Anand, V., Biondich, P. G., & Carroll, A. E. (2008). The CHICA smoking cessation system. In AMIA Annual Symposium Proceedings (Vol. 2008, p. 166). American Medical Informatics Association.
15. Downs, S. M., Marasigan, F., Abraham, V., Wildemuth, B., & Friedman, C. P. (1999). Scoring performance on computer-based patient simulations: beyond value of information. In Proceedings of the AMIA Symposium (p. 520). American Medical Informatics Association.
16. Anand, V., & Downs, S. M. (2008). Probabilistic asthma case finding: a noisy or reformulation. In AMIA Annual Symposium Proceedings (Vol. 2008, p. 6). American Medical Informatics Association.
17. * Klann, J. G., Anand, V., & Downs, S. M. (2013). Patient-tailored prioritization for a pediatric care decision support system through machine learning. Journal of the American Medical Informatics Association, 20(e2), e267-e274.
18. * Anand, Vibha, Aaron E. Carroll, Paul G. Biondich, Tamara M. Dugan, and Stephen M. Downs. "Pediatric decision support using adapted arden syntax."*Artificial Intelligence in Medicine* (2015).
