## Supplemental File and Appendix for "Designing a Clerkship Curriculum for Medical Students in Clinical and Medical Informatics in the Electronic Medical Record Era": Programming (with Python) and Databases - Pages Concatenated.docx

### Introduction: Programming

Work through the book Practical Programming: An Introduction to Computer Science Using Python 3

Exercises from this book will be used as assignments.

We recommend the [SciTE editor](http://www.scintilla.org/SciTEDownload.html) to create your assignments (the .py format files containing python code).

Python can be found at <https://www.python.org/>, and the standard distribution comes with An Integrated DeveLopment Environment for Python (IDLE), in which you can write dynamic code in a python show, as well as new .py python files, and run those files (the shortcut is F5) from the same set of windows.

The [Python For Beginners Guide](https://www.python.org/about/gettingstarted/) can also be found on the python.org mainpage.

### Assignment: Practical Programming Exercises 2.10

Upload answers to problems: 3, 5.

### Assignment: Practical Programming Exercises 3.11

Upload answers to problems: 5, 7, 10

### Assignment: Practical Programming Exercises 4.7

Upload answers to problems: 2, 4, 8, 9.

### Assignment: Practical Programming Exercises 5.6

Upload answers to problems: 4, 5, 8

### Assignment: Practical Programming Exercises 6.6

Upload answers to problems: 1, 2, 3

### Assignment: Practical Programming Exercises 7.6

Upload answers to problems: 10, 11, 12

### Assignment: Practical Programming Exercises 8.9

Upload answers to problems: 4, 5, 6, 8, 10, 11

### Assignment: Practical Programming Exercises 9.10

Upload answers to problems: 4, 5, 8, 13, 16

### Assignment: Practical Programming Exercises 10.10

Upload answers to problems: 2, 5, 6, 7, 8

### Assignment: Practical Programming Exercises 11.8

Upload answers to problems: 3, 8, 9, 11

### Assignment: Practical Programming Exercises 12.4

Upload answers to problems: 1, 3, 6

### Assignment: Practical Programming Exercises 14.8

Upload answers to problems:1, 2, 5

### Assignment: Practical Programming Exercises 17.10

Upload answers to problems: 1, 3

### Introduction: Databases

For this module you will be completing each of the mini-courses in the Stanford Self-Paced Database Course:

<https://lagunita.stanford.edu/courses/DB/2014/SelfPaced/about>

For each assignment, your task is to upload the statement of completion for each mini-course.

The order in which the courses is presented following this page is the suggested order in which one might find a logical flow to the material.

Jennifer Widom's reference text for introductory database course material (Database Systems: The Complete Book) can be found in the library, should you require additional source material (including from their book homepage <<http://infolab.stanford.edu/~ullman/dscb.html>>).

### Assignment: Relational Algebra

Pertinent Chapters: 2 and 4

- Section 2.3 Exercise 2.3.1
- Section 2.4 Exercises 2.4.1, 2.4.2, 2.4.5
- Section 2.5 Exercise 2.5.1
- Section 4.1 Exercises 4.1.1, 4.1.2, 4.1.6, 4.1.7, 4.1.8
- Section 4.4 Exercise 4.4.3

### Assignment: Unified Modeling Language

Pertinent Chapters: 4, 7 and 8

- Section 4.2 Exercises 4.2.5, 4.2.6, 4.2.7
- Section 4.5 Exercises 4.5.4
- Section 4.6 Exercise 4.6.3
- Section 4.7 Exercises 4.7.1,4.7.2, 4.7.4, 4.7.6, 4.7.9
- Section 4.8 Exercise 4.8.1

### Assignment: Relational Design Theory

Pertinent Chapter: 3

- Section 3.3 Exercise 3.3.1
- Section 3.4 Exercise 3.4.1
- Section 3.5 Exercises 3.5.1,3.5.2
- Section 3.6 Exercises 3.6.2, 3.6.3

### Assignment: Indexes and Transactions

Pertinent Chapters: 6 and 8

- Section 6.3 Exercises 6.3.1, 6.3.3, 6.3.8
- Section 6.4 Exercises 6.4.1, 6.4.6
- Section 6.5 Exercise 6.5.1
- Section 6.6 Exercises 6.6.1, 6.6.2, 6.6.3
- Section 8.3 Exercise 8.3.1
- Section 8.4 Exercises 8.4.1, 8.4.2

### Assignment: Constraints and Triggers

Pertinent Chapter: 7

- Section 7.1 Exercise 7.1.3
- Section 7.2 Exercise 7.2.2
- Section 7.3 Exercise 7.3.1
- Section 7.4 Exercise 7.4.1
- Section 7.5 Exercise 7.5.2

### Assignment: SQL

Pertinent Chapters: 5 and 6

- Section 5.1 Exercises 5.1.1, 5.1.2
- Section 5.2 Exercise 5.2.1
- Section 5.3 Exercise 5.3.1
- Section 5.4 Exercises 5.4.1, 5.4.2, 5.4.5
- Section 6.1 Exercises 6.1.3, 6.1.5
- Section 6.2 Exercises 6.2.2, 6.2.4, 6.2.5

### Assignment: Recursion in SQL

Pertinent Chapter: 10

- Section 10.2 Exercise 10.2.1
- Section 10.3 Exercises 10.3.1, 10.3.2, 10.3.3
- Section 10.4 Exercises 10.4.1, 10.4.2, 10.4.3
- Section 10.5 Exercises 10.5.1, 10.5.2
