## Supplemental File and Appendix for "Designing a Clerkship Curriculum for Medical Students in Clinical and Medical Informatics in the Electronic Medical Record Era": Terminologies and Standards - Pages Concatenated.docx

### The UMLS: Metathesaurus et al

The Unified Medical Language System (UMLS) is a software system that integrates information from both clinical and biomedical terminologies/standards. Started in 1986, and first described by *Humphreys et al*, it serves as a long term research and development project of the National Library of Medicine, with researchers at the [Lister Hill National Center for Biomedical Communications](http://lhncbc.nlm.nih.gov/) serving its aims. As described by the NLM:

the UMLS project is an effort to overcome two significant barriers to effective retrieval of machine-readable information. The first is the variety of ways the same concepts are expressed in different machine-readable sources and by different people. The second is the distribution of useful information among many disparate databases and systems

[The UMLS Reference Manual](http://www.ncbi.nlm.nih.gov/books/NBK9676/) describes the technical features of the knowledge sources, software and related services. The UMLS has three key tools:

1. [Metathesaurus](http://www.nlm.nih.gov/research/umls/knowledge_sources/metathesaurus/index.html): The metathesaurus is the mapping project which links terms and codes from vocabularies together. [Appendix 1](http://www.nlm.nih.gov/research/umls/knowledge_sources/metathesaurus/release/license_agreement_appendix.html) to the UMLS Metathesaurus license agreement contains a list of all of the source vocabularies.
2. [Semantic Network](http://semanticnetwork.nlm.nih.gov/SemGroups/): The semantic network serves as a mechanism to categorize the concepts represented by the metathesaurus, described by *McCray et al*. It also serves to link concepts and groups together through semantic relationships. The network groups the 99.5% of the metathesaurus into 15 semantic groups:
   1. Activities & Behaviors
   2. Anatomy
   3. Chemicals & Drugs
   4. Concepts & Ideas
   5. Devices
   6. Disorders
   7. Genes & Molecular Sequences
   8. Geographic Areas
   9. Living Beings
   10. Objects
   11. Occupations
   12. Organizations
   13. Phenomena
   14. Physiology
   15. Procedures
2. SPECIALIST Lexicon and Lexical Tools: The specialist lexicon and tools, developed by scientists at Lister Hill, serve to provide natural language processing capability and mediate between clinical text and the other resources of the UMLS. Specifically these tools include:
   1. [SPECIALIST Lexicon](http://lexsrv3.nlm.nih.gov/Specialist/Summary/lexicon.html)
   2. [LexAccess](http://lexsrv3.nlm.nih.gov/Specialist/Summary/lexAccess.html)
   3. [Lexical Tools](http://lexsrv3.nlm.nih.gov/Specialist/Summary/lexicalTools.html)
   4. [Text Tools](http://lexsrv3.nlm.nih.gov/Specialist/Summary/textTools.html)
   5. [Text Categorization](http://lexsrv3.nlm.nih.gov/Specialist/Summary/textCategorization.html)
   6. [GSpell](http://lexsrv3.nlm.nih.gov/Specialist/Summary/gSpell.html)
   7. [DTagger](http://lexsrv3.nlm.nih.gov/Specialist/Summary/dTagger.html)
   8. [Visual Tagging Tool](http://lexsrv3.nlm.nih.gov/Specialist/Summary/vtt.html)
   9. [Sub-Term Mapping Tools](http://lexsrv3.nlm.nih.gov/Specialist/Summary/stmt.html)
   10. [MEDLINE N-Gram](http://lexsrv3.nlm.nih.gov/LexSysGroup/Projects/nGram/index.html) These tools provide for both the translation from biomedical and general English to UMLS terms/the SPECIALIST lexicon itself, but also provide Java-based software for parsing, tagging, annotation and other computational linguistic techniques.

### UMLS Terminology Services

[The UMLS Terminology Services](https://uts.nlm.nih.gov/home.html) is the hub for UMLS services and tools. It allows for access to both local and web-hosted tools. Specifically described as contained in the services package are the ability to:

1. Administrative actions
   - Request a UMLS Metathesaurus/UTS account
   - Report on use of UMLS and its services
2. Use web-based applications:
   - Metathesaurus Browser
   - Semantic Network Browser
   - SNOMED CT Browser
3. Download UMLS Knowledge sources, updates
4. Query web-services via the Application Program Interface (API)

The UTS offers the core web-based applications listed above, as well as other NLM applications which link into the UMLS:

- [MetaMap](http://metamap.nlm.nih.gov/) is a tool developed to map biomedical text to the UMLS Metathesaurus
- [MyMedicationList](http://mml.nlm.nih.gov/) is a tool to reconcile personal medication records and medication history, as described by *Nelson, Zeng and Bodenreider*.
- [RxNav](http://mor.nlm.nih.gov/download/rxnav/) is a browser for drug information services and standards to allow for cross linkages, as well as clinical information presentation.
- [RxTerms](https://wwwcf.nlm.nih.gov/umlslicense/rxtermApp/rxTerm.cfm) is a publicly available interface for prescription writing and/or medication history recording.
- [DailyMed](http://dailymed.nlm.nih.gov/dailymed/) is an application that provides information collected from FDA labels and package inserts for marketed drugs, both human and animal.
- [Semantic Navigator](http://morc1.nlm.nih.gov:8000/perl/auth/semnav.pl) is a navigator to examine the semantic relationships between terms identified by both Concept Unique Identifier (CUI) and plain English.
- Tools from the SPECIALIST lexicon and lexical tools: LexAccess, Lexical Tools
- [MedlinePlus Connect](http://www.nlm.nih.gov/medlineplus/connect/overview.html), which provides a portal connect electronic medical records to [MedlinePlus](http://www.nlm.nih.gov/medlineplus/), a curated consumer-facing health information resource. This connection supports the use of UMLS terminologies as a means to search for information, rather than the standard plain language interface of MedlinePLus.

##### UTS API v2.0

The [documentation](https://uts.nlm.nih.gov//home.html#apidocumentation) for the API describes the entirety of the methods available to access UMLS services, which are configured to operate via the Simple Object Access Protocol (SOAP), by exchanging files formatted in eXtensible Markup Language (XML). The categories of methods within the API are:

1. Paging, Sorting and Filtering: allows for the customization of web service calls in a variety of ways to include or exclude content.
2. Metathesaurus Content: allows for the querying and examining of the metathesaurus and examination of particular content (terms) and hierarchies.
3. Metathesaurus Metadata: allows for the accessing of UMLS metadata surrounding sources, entries, terms and relationships…
4. Semantic Types: allows for the querying of the semantic network and the determination of the relationships between entities within the network.
5. Concept History: allows for the querying of the history of the UMLS metathesaurus, and temporal trends related to concepts.
6. Finder Service: allows for the retrieving of identifiers and exchange them for other concepts at varying levels of granularity, allowing for the transposition of encoded data.

##### UTS API Example

The following code uses the [SUDS](https://fedorahosted.org/suds/) module, a soap-based client for web services for Python, to access the UTS API.

```
#import Client function from suds.client module
from suds.client import Client
#import both the getpass module and suds module
import getpass, suds
#url for security client
security_url = 'https://uts-ws.nlm.nih.gov/services/nwsSecurity?wsdl'
#pass url for security client to Client function to obtain object
security_client = Client(security_url)
print("UMLS UTS Login")
#get login username from standard input using raw_input function
user_name = raw_input('Username: ')
#get password using the getpass module (returns seKr3t input with echo turned off, as to prevent over the shoulder attack)
password = getpass.getpass('Password: ')
#error checking for incorrect password/username
#proxy grant ticket
#attempt to obtain proxy_ticket, if failure (due to incorrect password) exit()
try:
    proxy_ticket = security_client.service.getProxyGrantTicket(user_name,password)
except suds.WebFault:
    print("Incorrect UMLS UTS username or password")
    exit()
#run client side query of web service
#set content url
content_url = 'https://uts-ws.nlm.nih.gov/services/nwsContent?wsdl'
#get client object from Client function for content url
content_client = Client(content_url)
#generate a single use ticket using the web security_client.service getProxyTicket function
single_use_ticket = security_client.service.getProxyTicket(proxy_ticket,'http://umlsks.nlm.nih.gov')
#using the content_client.service getCode function, pass in the single use ticket obtained from the security client service, as well as the umls version, term and source in order to obtain an object in return
concept_api = content_client.service.getCode(single_use_ticket,umls_version,umls_term,umls_source)
#print the concept out to standard output
print(concept_api)
```

### SNOMED-CT

From 20 years prior to 1993 and since, the College of American Pathologists (CAP) developed The Systematized Nomenclature of Human and Veterinary Medicine International (SNOMED-I). This terminology standard led to the development of the SNOMED Reference Terminology (SNOMED-RT). SNOMED-RT served as a reference terminology to complement the medical concepts in SNOMED-I, representing multiple hierarchies and decision logic to provide multi-level granularity to the operations of health care. Combining this terminology with the United Kingdom’s National Health Service’s Clinical Terms (Read Codes) formed the beginnings of SNOMED-CT (SNOMED - Clinical Terms). SNOMED-CT subsequently has evolved into the most comprehensive English language clinical vocabulary, and serves as the Federal standard for interoperability. SNOMED-CT is currently owned by the International Health Terminology Standards Organisation (IHTSDO), after purchasing the intellectual property from the CAP in 2007. The National Library of Medicine pays an annual fee (to the order of $6 million), in order to make SNOMED-CT available to anyone in the United States. As you are undoubtedly aware, the license for the UMLS Metathesaurus includes a section regarding SNOMED-CT, which describes is use and operation under license.

Rather than codes, SNOMED-CT is primarily about terms and their relationships. An example of the terms and relationships surrounding the English term “Heart Attack” courtesy of the Semantic Navigator tool follows:

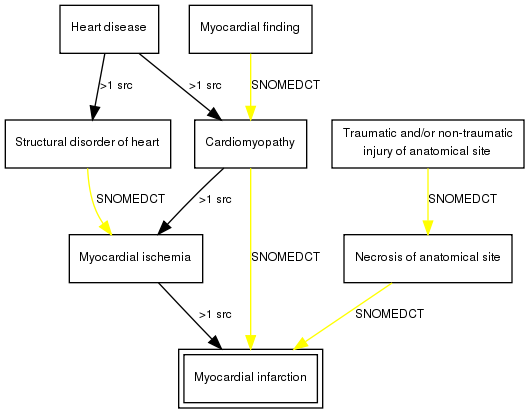

The relationships between concepts due to SNOMED-CT can be seen via the yellow lines, as well as the black *>1 src* linkages. By using the [Semantic Navigator](http://mor3.nlm.nih.gov:8000/perl/auth/semnav.pl), and restricting to the SNOMED-CT vocabulary, one can explore terms dynamically (see in-line frame below).

The hierarchies of SNOMED CT begin with the following 19 categories, and subsequently drill down as can be seen in the UTS [Metathesaurus Browser’s](https://uts.nlm.nih.gov//metathesaurus.html) tree view (see in-line frame below).

1. Body structure
2. Clinical finding
3. Environment or geographical location
4. Event
5. Observable entity
6. Organism
7. Pharmaceutical/biologic product
8. Physical force
9. Physical object
10. Procedure
11. Qualifier value
12. Record artifact
13. SNOMED CT Model Component
14. Situation with explicit context
15. Social context
16. Special concept
17. Specimen
18. Staging and scales
19. Substance

SNOMED-CT has been shown to be able to model or serve as a terminological replacement to the problem list by *Elkin et al*. Such a replacement could allow for more effective relationships be explored between conditions due to known ontological definitions, as well as driving decision support systems that use the problem list to trigger rules (such as to identify patients with specific diseases or phenotypes). SNOMED-CT also supports the identification of concepts within clinical text and serves as a machine readable method to examine clinical semantics.

### ICD-9/ICD-10

The International Classification of Disease (ICD) versions 9 and 10 are standards maintained by the [World Health Organization](http://www.who.int/classifications/icd/en/). It particularly functions to identify disease related to morbidity and mortality for the comparison at the international level. This standard is argued to have originally built off of François Bossier de Lacroix’s *Nosologia methodica sistens morborum classes, genera et species, juxtà sydenhami mentem & botanicorum ordinem*, the man credited as first in the nosological classification of disease. This evolved into William Cullen’s *Synopsis Nosologiae Methodicae* in Edinburgh. From this attempt at classification, observations upon the London Bills of Mortality were written by John Graunt. There is some dispute as to the authorship and the nature of these bills, as described by C H Hull in his work *Graunt or Petty? The Authorship of the Observations Upon the Bills of Mortality* John Graunt’s observations as well as the aforementioned bills can be found in the collected work entitled *A Collection of the Yearly Bills of Mortality from 1657 to 1758 inclusive together with several other bills of an earlier date*. This classification served as the precursor to the ICD, as a means to expand Cullen’s work on the nological classification of medicine and incorporate it for demographic and statistical purposes for the Registrar General of England, under the direction of William Farr. In this report, Farr opined that:

The advantages of a uniform statistical nomenclature, however imperfect, are so obvious, that it is surprising no attention has been paid to its enforcement in Bills of Mortality. Each disease has, in many instances, been denoted by three or four terms, and each term has been applied to as many different diseases: vague, inconvenient names have been employed, or complications have been registered inst ead of primary diseases. The nomenclature is of as much importance in this department of inquiry as weights and measures in the physical sciences, and should be settled without delay.

The ICD itself grew out of the meetings of the International Statistical Congress, as well as its successor the International Statistical Institute. In 1891, the Bertillion Classification of Causes of Death was created, named after Jacques Bertillion, the Chief of Statistical Services of the City of Paris. This classification drew on French, English, German and Swiss classifications, and was built on Farr’s principles. The American Public Health Association in 1898 recommended the adoption of this classification for Canada, Mexico and the United States. Following the International Statistical Institute’s conference in 1899, where Bertillion’s revisions among others were reported, the French Government called the first International Conference for the Revision of the Bertillion or International List of Causes of Death. From then on, in ten year increments, this classification evolved, until the death of Bertillion in 1922. Subsequently to Bertillion, with the support of the Health Organization of the League of Nations (a precursor to the World Health Organization under the United Nations), the fourth and fifth revisions of the International List of Causes of Death were developed. Following the second decennial revision, the United States Department of Commerce and Labour’s translation, re-purposed for mortality statistics was named *International Classification of Causes of Sickness and Death*. This, alongside the Standard Morbidity Code developed by the Dominion Council of Health of Canada in 1936, served to further push the now nascent ICD into the realm of mortality statistics (as seen valuable by state statistical and census-taking organizations). This sixth decennial revision in 1946 formally adopted the name for the newly focused classification of *International Classification of Diseases, Injuries, and Causes of Death*, and the WHO was formally involved (and endorsed) the subsequent report of the sixth revision’s conference of 1948. This increased the international viability of the classification, and focused it more heavily on performing in the mortality statistics arena. With the seventh and eighth revisions, the use of ICD for indexing medical records, as well as the national specificity (via country-specific adaptations) was enhanced. Following this, the ninth revision was made in 1975, and its localization for the United States remains in place today. A further, more comprehensive history of the ICD can be found in the report of Centers for Disease Control and Prevention’s National Center for Health Statistics entitled *History of the Statistical Classification of Diseases and Causes of Death*.

The tenth revision was released in 1994, and endorsed by the WHO (and is required in the US for death statistics). It offers a more specific and effective ontology than the ninth revision, and is slated to roll out to replace ICD-9 in October 2015 (although that compliance date has already been pushed back a year previously, and lobbying efforts may introduce further delays). An example of such lobbying is that of the American Medical Association, which has advocated for a variety of terms and conditions. Their list of frequently asked questions and a detailed discussion of the circumstances involved in the replacement can be found [here](http://www.ama-assn.org/ama/pub/physician-resources/solutions-managing-your-practice/coding-billing-insurance/hipaahealth-insurance-portability-accountability-act/transaction-code-set-standards/icd10-code-set.page). In discussing the subsequent changes, Farr’s opinion remains relevant today, as concerns over the value of the specificity of new revisions and codes have been made (particularly within the United States).

Unlike SNOMED-CT, the ICD is a discrete coding system. The current revision of ICD-10, as maintained by the WHO (unlocalized to a specific country) can be found [here](http://apps.who.int/classifications/icd10/browse/2015/en) The ICD-11 Beta Draft can be found [here](http://apps.who.int/classifications/icd11/browse/l-m/en) The UMLS contains the ICD9 and ICD10 coding systems, both with clinical modification as well as some localizations for countries such as Australia. The code found accessing the UTS API can be used to identify specific codes, while the UMLS Metathesaurus Browser can be used to examine terms and their relationships (in both search and tree forms).

#### ICD 9th Revision, Clinical Modification (ICD-9 CM)

Consists of a three digit category, a period, then single digit subcategory and subclassification numbers. Three digits can be prepended by *V* for coding supplementary health status/health services contact (V01-V89), or *E* for external causes of injury and poison (E800-E999). You can use the UTS Metathesaurus Browser to view various releases of the ICD9CM codes within the context of the UMLS. Examples include:

- 410.0/Acute myocardial infarction, of anterolateral wall
- 097.0/Late syphilis, unspecified
- 277.0/Cystic fibrosis
- 277.02/Cystic fibrosis with pulmonary manifestations
- 125.1/Malayan filariasis

Beyond diagnoses, the ICD-9 CM also contains coding for procedures. These align with ICD-10 PCS codes, or [Current Procedural Terminology codes](http://www.ama-assn.org/ama/pub/physician-resources/solutions-managing-your-practice/coding-billing-insurance/cpt.page), a codeset run out of the American Medical Association to report procedures and services. For example:

- 42.4/Excision of esophagus
- 88.71/Diagnostic ultrasound of head and neck

#### ICD 10th Revision, Clinical Modification (ICD-10 CM)

The code is blocked into segments, separated by periods at the three and six character mark. It offers more specificity than ICD-9 CM, and supports more accurate classification (and is currently in use with death statistics across the world):

1. The first character being a letter (A-T) associated with a (part of a) chapter (or two) detailing the core hierarchy. U codes are used for the WHO for provisional assignment (U00-U49) and research (U50-U99):

Chapters I-XVII: diseases and other morbid conditions Chapter XIX: injuries, poisoning and certain other consequences of external causes Chapter XVIII: Symptoms, signs and abnormal clinical and laboratory findings, not elsewhere classified Chapter XX: External causes of morbidity and mortality Chapter XXI: Factors influencing health status and contact with health services

2. A following multi-character segment consisting of one or more of:
   1. Three character segment reflecting two axes of classification within the chapter identified by the first character
   2. Fourth character segment to delineate divisions within the three character category or groups of conditions
   3. Fifth character segment to delineate further divisions from the fourth character category

Examples include:

- I21.0/Acute transmural myocardial infarction of anterior wall
- A52.9/Late syphilis, unspecified
- E84/Cystic fibrosis
- E84.0/Cystic fibrosis with pulmonary manifestations
- B74.1/Filariasis due to Brugia malayi

#### ICD 10th Revision, Procedure Coding System (ICD-10 PCS)

The ICD-10 PCS is a system consisting of seven character codes, each character illustrating a particular classification hierarchy. These codes describe procedures classified within sub-categories by nested hierarchies.

1. Section
2. Body System
3. Root Operation
4. Body Part
5. Approach
6. Device
7. Qualifier

Examples include:

- U002491/Oesophagus: Excision
- BH4CZZZ/Ultrasonography of Head and Neck

### RxNorm

RxNorm is a terminology system to normalize names of clinical drugs and supporting semantic operation between drug information and pharmacy systems. It specifically operates for both generic and branded:

- Pharmaceutical products provided to or consumed by a patient with a purpose related to therapeutics or diagnostics
- Sets containing multiple drugs, or drugs designed to be administered in a specified sequence, not including radiopharmaceuticals, bulk powders, contrast media, food, or dietary supplements

Out of the standards presented, RxNorm is the youngest standard, having been initially born out of an experiment in modeling clinical drugs in the UMLS in 2001. The project subsequently evolved, with the assistant of HL7’s vocabulary technical committee, and documentation for the entirety of the terminology standard can be found [here](http://www.nlm.nih.gov/research/umls/rxnorm/docs/2015/rxnorm_doco_full_2015-1.html) RxNorm is a mechanism to synthesize drug standards from a variety of source vocabularies, a table of these can be found in section 3.1 of the technical documentation. Like the UMLS Metathesaurus, the true form of RxNorm is as a series of interrelated concepts. It serves to be able to link concepts related to the use, delivery and operationalization of drugs in a standardized fashion, capable of linking between pharmacies, FDA descriptions/information sources and hospitals.

In order to view RxNorm, [RxNav](https://rxnav.nlm.nih.gov/index.html) is a browser for drug information sources, including RxNorm, with a dedicated [API for RxNorm](https://rxnav.nlm.nih.gov/RxNormAPIs.html). Figure 1 from*Nelson et al* describes the relationships linking a single drug *Cetirizine Hydrochloride (Zyrtec/Cetirizine)* with the concepts within RxNorm (reproduced below).

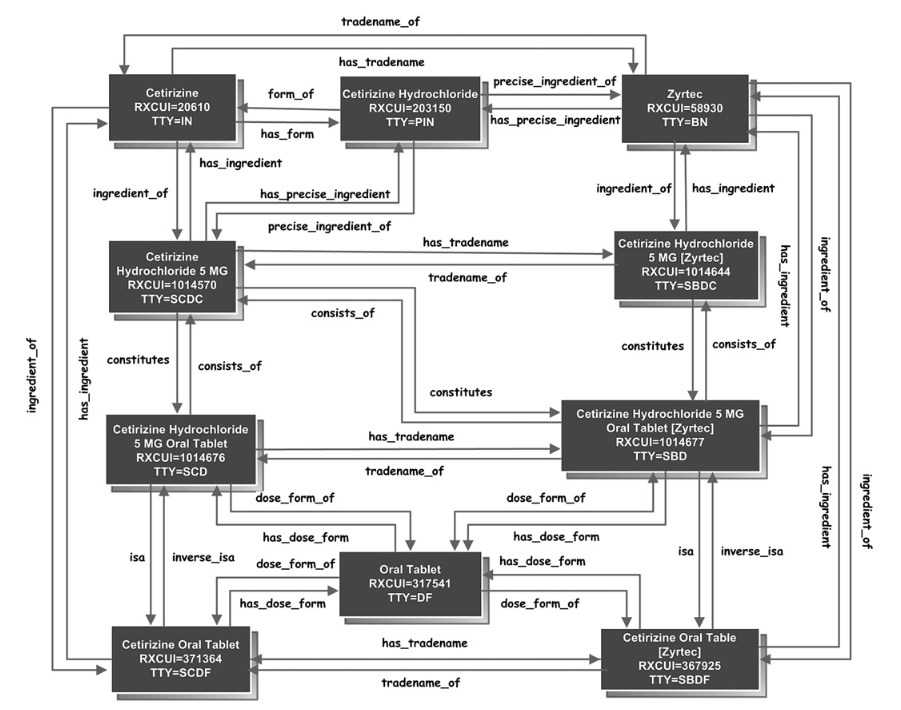

### HL7 v2.8 and FHIR

#### [Create a HL7 Account](http://qcommerce.hl7.org/qcommercenet/NewUser/Index.aspx)

[Health Level Seven International](http://www.hl7.org/index.cfm) is a standards organization that manages several standards, the master grid of which can be found [here](http://www.hl7.org/implement/standards/product_matrix.cfm?ref=nav). These standards include:

1. Arden Syntax, a standard for describing logic in clinical circumstances
2. The HL7 messaging standard, for communication between organizations, devices and other sources
3. The Fast Healthcare Interoperability Resources Specification (FHIR)

#### Arden Syntax

Arden Syntax originated from a meeting in 1989 at the Arden Homestead in New York. Following this meeting, American Society for Testing and Materials published the first specification in 1992. Incrementally, Arden Syntax has been subsequently iterated, and is currently at version 2.10. In 1999, HL7 took over publication of Arden Syntax with version 2.0 in 1999.

Arden Syntax can be described as a programming language specifically focused on the implementation of clinical decision support. *Hripcsak et al*, describes the initial rationale and methods for writing Arden syntax in his seminal 1994 articles. Describing version 2.8, *Samwald et al*describes the particulars of Arden Syntax’s operation, and its value as a hybrid between:

1. Classical production rules
2. Procedural representation of clinical algorithms

The core of Arden Syntax is the medical logic module, a block of code that describes the logic to be activated by information provided by the clinical information system, or procedurally from clinical operations. Examples can be found in the *Samwald et al* paper, as well as the Arden Syntax version 2.9 Manual Section X4.

#### HL7 Messaging and FHIR

Work on the version 1.0 draft standard was first begun in 1987, initially describing interfaces, admission/discharge/transfer, order entry and display-oriented queries. This draft was presented on October 8th 1987. Version 2.0 was presented in September 1988, the second plenary since the initial draft 1987. Further iteration has occurred since, with major milestones at versions 2.1, 2.2, 2.3, 2.3.1, 2.4, 2.5, 2.5.1, 2.6, 2.7, 2.8, and now 2.8.1.

HL7 messaging is “*the workhorse of electronic data exchange in the clinical domain*” according the [HL7 V2.x Product Brief](http://www.hl7.org/implement/standards/product_brief.cfm?product_id=185). The version 2.x standards date back to 1989, and currently stand at version 2.8.1. It serves to encode information in a meaningful way capable of interfacing between various systems:

- sending or receiving patient admissions/registration, discharge or transfer (ADT) data
- sending or receiving queries, resource and patient scheduling orders
- results
- clinical observations
- billing
- master file update information
- medical records
- scheduling
- patient referral
- patient care
- clinical laboratory automation
- application management
- personnel management messages

The HL7 messaging system describes its encoding rules in the Health Level Seven, Version 2.8.1 Final Standard:

Data fields that are of variable length and separated by a field separator character. Rules describe how the various data types are encoded within a field and when an individual field may be repeated. The data fields are combined into logical groupings called segments. Segments are separated by segment separator characters. Each segment begins with a three-character literal value that identifies it within a message. Segments may be defined as required or optional and may be permitted to repeat. Individual data fields are found in the message by their position within their associated segments.

All data is represented as displayable characters from a selected character set. The ASCII displayable character set (hexadecimal values between 20 and 7E, inclusive) is the default character set unless modified in the MSH header segment. The field separator is required to be chosen from the ASCII displayable character set. All the other special separators and other special characters are also displayable characters, except that the segment separator is the ASCII Carriage Return character.

The encoding rules distinguish between data fields that have the null value and those that are not present. The former are represented by two adjacent quotation marks, the latter by no data at all (i.e., two consecutive separator characters.) The distinction between null values and those that are not present is important when a record is being updated. In the former case the field in the database should be set to null; in the latter case it should retain its prior value. The encoding rules specify that if a receiving application cannot deal with a data field not being present, it should treat the data field as present but null.

The encoding rules specify that a receiving application should ignore fields that are present in the message but were not expected rather than treat such a circumstance as an error. For more information on fields and encoding rules, see Section 2.5.3, “Fields,” and 2.6, “Message Construction Rules.”

The details of the standard along with examples can be found in hierarchical form using the UMLS Metathesaurus Browser, or by reading the standard. In order to download the standard, you must create an account on HL7’s website using the link at the top of this page.

Since the development of HL7 v2.7/2.8, those working on the messaging standard have been building to advance the next generation of the messaging standard. The HL7 Version 3 [normative standard](http://www.hl7.org/implement/standards/product_brief.cfm?product_id=186) was introduced as an intended replacement for the HL7 v2.x standard. Specifically, it uses the eXtensible Markup Language (XML) to describe domains which provide use-case focused storyboard descriptions, trigger events, interaction designs, domain object models as a means of encoding the message around HL7’s [Reference Information Model](http://www.hl7.org/implement/standards/rim.cfm). It has inadequately been taken up in the United States, and although reports use within the United Kingdom’s National Health Service, Dutch and Canadian national infrastructure blueprints, it is intended to be replaced by FHIR. [Fast Healthcare Interoperability Resources (FHIR)](http://www.hl7.org/fhir/overview.html) is the latest specification intending to encode and exchange data. It still retains the use-case ideology of HL7 v3, however provides extensibility through APIs and XML and JSON formats. A more formal comparison between FHIR and the other messaging standards (v2 and v3) can be found [here](http://www.hl7.org/fhir/comparison.html). It is currently in version 0.0.82, and progressing along as a draft standard for trial use.

### LOINC

Logical Observation Identifiers Names and Codes (LOINC) is a standard used for encoding clinical observations. A great deal of these observations include laboratory tests and orders. Dan Vreeman describes LOINC as:

If an observation is a question and the obsevation value is an answer… LOINC provides codes for*questions*, other vocabularies provide codes for *answers*

You can find the complete table and user guide for LOINC [here](http://loinc.org/downloads/loinc). LOINC codes are described along six axes:

1. Component: what is being measured or observed, the “test”
2. Property: characteristic of what is being measured
3. Timing: interval in time in which observation is made
4. Sample (or system): the type of sample or system
5. Scale: quantitative/ordinal/nominal/narrative
6. Method (if indicated): method or procedure used to produce the result or observation

Timing and system/sample can be divided into two subparts: max,min,mean and source when not the patient. The [Regenstrief LOINC Mapping Assistant](http://loinc.org/downloads/relma) can be used to search for LOINC terms using plain language, as well as map to a local terminology set. Along with the UMLS Metathesaurus, this can be used to explore the standard. The Regenstrief Institute (through Clem McDonald) has led the stewardship of the laboratory side of the LOINC codes, with Stanley Huff of Intermountain Healthcare serving on the pathological/clinical side.

### Assignment: Integrating Terminologies for Diagnosis/Disease Classification

1. Select your favourite disease.
2. Using the UTS, and other resources described, detail how this disease is described by multiple standards, along with the rationale.
3. Describe how these standards interact (or not) to form a complete picture of disease.
4. Submit a report with the above content, that describes how the terminology standards which surround the disease (and its diagnosis), impact the concept of the disease, as well as potential clinical management.
