## Supplemental File and Appendix for "Designing a Clerkship Curriculum for Medical Students in Clinical and Medical Informatics in the Electronic Medical Record Era": Pre-Course_Questionnaire.html

Quiz: Pre-Course Questionnaire


### You need to have JavaScript enabled in order to access this site.


Close

##### Loading...

### Pre-Course Questionnaire

#### Quiz Instructions

This is the pre-course questionnaire, please fill this out so we can get to know you prior to you joining us in the course.


2019-01-30T20:55:32-05:00
2019-01-30T20:55:33-05:00
2060-06-29T22:59:00-05:00

 
1306893807

*Move To...

This element is a more accessible alternative to drag & drop reordering. Press Enter or Space to move this question.*

Flag this Question

Question 1

0 pts

*Edit this Question*
*Delete this Question*

0
fill\_in\_multiple\_blanks\_question

What is your name (Firstname,Lastname): <input
class='question\_input'
type='text'
autocomplete='off'
style='width: 120px;'
name='question\_105589\_3cb984d64966581c40dc5ba9713ca2b3'
value='{{question\_105589\_3cb984d64966581c40dc5ba9713ca2b3}}' />
, <input
class='question\_input'
type='text'
autocomplete='off'
style='width: 120px;'
name='question\_105589\_61df38c0a2ed6f26e8fd64c4f122576f'
value='{{question\_105589\_61df38c0a2ed6f26e8fd64c4f122576f}}' />

What is your name (Firstname,Lastname): 
,

*Move To...

This element is a more accessible alternative to drag & drop reordering. Press Enter or Space to move this question.*

Flag this Question

Question 2

0 pts

*Edit this Question*
*Delete this Question*

0
multiple\_choice\_question

With what gender do you identify?

With what gender do you identify?

|  |  |
| --- | --- |
|  | Male |

|  |  |
| --- | --- |
|  | Female |

|  |  |
| --- | --- |
|  | Other |

*Move To...

This element is a more accessible alternative to drag & drop reordering. Press Enter or Space to move this question.*

Flag this Question

Question 3

0 pts

*Edit this Question*
*Delete this Question*

0
numerical\_question

What is your age?

What is your age?

*Move To...

This element is a more accessible alternative to drag & drop reordering. Press Enter or Space to move this question.*

Flag this Question

Question 4

0 pts

*Edit this Question*
*Delete this Question*

0
short\_answer\_question

What is your current degree program: [degree]

What is your current degree program: [degree]

*Move To...

This element is a more accessible alternative to drag & drop reordering. Press Enter or Space to move this question.*

Flag this Question

Question 5

0 pts

*Edit this Question*
*Delete this Question*

0
multiple\_choice\_question

 
256009

Do you have a particular interest in a clinical specialty (due to research, or future or current practice)?

Do you have a particular interest in a clinical specialty (due to research, or future or current practice)?

|  |  |
| --- | --- |
|  | Anesthesiology |

|  |  |
| --- | --- |
|  | Dermatology |

|  |  |
| --- | --- |
|  | Emergency Medicine |

|  |  |
| --- | --- |
|  | Family Medicine |

|  |  |
| --- | --- |
|  | Internal Medicine |

|  |  |
| --- | --- |
|  | Med-Peds (Internal Medicine / Pediatrics) |

|  |  |
| --- | --- |
|  | Neurosurgery |

|  |  |
| --- | --- |
|  | Neurology |

|  |  |
| --- | --- |
|  | Obstetrics & Gynecology |

|  |  |
| --- | --- |
|  | Ophthalmology |

|  |  |
| --- | --- |
|  | Orthopedic Surgery |

|  |  |
| --- | --- |
|  | Otolaryngology (Ears, Nose, and Throat) |

|  |  |
| --- | --- |
|  | Pathology |

|  |  |
| --- | --- |
|  | Pediatrics |

|  |  |
| --- | --- |
|  | Physical Medicine & Rehabilitation |

|  |  |
| --- | --- |
|  | Plastic Surgery |

|  |  |
| --- | --- |
|  | Psychiatry |

|  |  |
| --- | --- |
|  | Radiation Oncology |

|  |  |
| --- | --- |
|  | Radiology |

|  |  |
| --- | --- |
|  | Surgery |

|  |  |
| --- | --- |
|  | Urology |

*Move To...

This element is a more accessible alternative to drag & drop reordering. Press Enter or Space to move this question.*

Flag this Question

Question 6

0 pts

*Edit this Question*
*Delete this Question*

0
true\_false\_question

 
256011

Have you previously participated in any research projects with a university or hospital environment?

Have you previously participated in any research projects with a university or hospital environment?

|  |  |
| --- | --- |
|  | True |

|  |  |
| --- | --- |
|  | False |

*Move To...

This element is a more accessible alternative to drag & drop reordering. Press Enter or Space to move this question.*

Flag this Question

Question 7

0 pts

*Edit this Question*
*Delete this Question*

0
multiple\_answers\_question

 
256020

Why did you select this elective? (You may select more than one)

Why did you select this elective? (You may select more than one)

|  |  |
| --- | --- |
|  | Previously described by former students as a bird course |

|  |  |
| --- | --- |
|  | Offers a connection to the Regenstrief Institute |

|  |  |
| --- | --- |
|  | Interest in data analytics |

|  |  |
| --- | --- |
|  | Interest in big data |

|  |  |
| --- | --- |
|  | Interest in medical administration |

*Move To...

This element is a more accessible alternative to drag & drop reordering. Press Enter or Space to move this question.*

Flag this Question

Question 8

0 pts

*Edit this Question*
*Delete this Question*

0
essay\_question

 
256019

Please describe why you decided to pursue this elective

Please describe why you decided to pursue this elective

HTML Editor
Rich Content Editor

*Move To...

This element is a more accessible alternative to drag & drop reordering. Press Enter or Space to move this question.*

Flag this Question

Question 9

0 pts

*Edit this Question*
*Delete this Question*

0
true\_false\_question

 
256005

Do you plan on an academic/research career?

Do you plan on an academic/research career?

|  |  |
| --- | --- |
|  | True |

|  |  |
| --- | --- |
|  | False |

*Move To...

This element is a more accessible alternative to drag & drop reordering. Press Enter or Space to move this question.*

Flag this Question

Question 10

0 pts

*Edit this Question*
*Delete this Question*

0
true\_false\_question

 
256013

Do you plan on using informatics in your future practice?

Do you plan on using informatics in your future practice?

|  |  |
| --- | --- |
|  | True |

|  |  |
| --- | --- |
|  | False |

*Move To...

This element is a more accessible alternative to drag & drop reordering. Press Enter or Space to move this question.*

Flag this Question

Question 11

0 pts

*Edit this Question*
*Delete this Question*

0
essay\_question

 
256015

What experience can we provide that would mean you find this elective worthwhile?

What experience can we provide that would mean you find this elective worthwhile?

HTML Editor
Rich Content Editor

*Move To...

This element is a more accessible alternative to drag & drop reordering. Press Enter or Space to move this question.*

Flag this Question

Question 12

0 pts

*Edit this Question*
*Delete this Question*

0
essay\_question

Define medical informatics

Define medical informatics

HTML Editor
Rich Content Editor

*Move To...

This element is a more accessible alternative to drag & drop reordering. Press Enter or Space to move this question.*

Flag this Question

Question 13

0 pts

*Edit this Question*
*Delete this Question*

0
essay\_question

Define biomedical informatics.

Define biomedical informatics.

HTML Editor
Rich Content Editor

*Move To...

This element is a more accessible alternative to drag & drop reordering. Press Enter or Space to move this question.*

Flag this Question

Question 14

0 pts

*Edit this Question*
*Delete this Question*

0
essay\_question

What problems do people in the field of medical informatics try to solve?

What problems do people in the field of medical informatics try to solve?

HTML Editor
Rich Content Editor

*Move To...

This element is a more accessible alternative to drag & drop reordering. Press Enter or Space to move this question.*

Flag this Question

Question 15

0 pts

*Edit this Question*
*Delete this Question*

0
essay\_question

How has the US government influenced the field of medical informatics?

How has the US government influenced the field of medical informatics?

HTML Editor
Rich Content Editor

*Move To...

This element is a more accessible alternative to drag & drop reordering. Press Enter or Space to move this question.*

Flag this Question

Question 16

0 pts

*Edit this Question*
*Delete this Question*

0
essay\_question

List some of the barriers to Health Information Technology (HIT) adoption.

List some of the barriers to Health Information Technology (HIT) adoption.

HTML Editor
Rich Content Editor

*Move To...

This element is a more accessible alternative to drag & drop reordering. Press Enter or Space to move this question.*

Flag this Question

Question 17

0 pts

*Edit this Question*
*Delete this Question*

0
essay\_question

Describe the educational and career opportunities within medical informatics.

Describe the educational and career opportunities within medical informatics.

HTML Editor
Rich Content Editor

Time's Up! Submitting results in:

Ok, fine

Not saved

Submit Quiz

a315800f-2a67-42ae-8f51-bd4f03e6b31d

 
