## Supplemental File and Appendix for "Designing a Clerkship Curriculum for Medical Students in Clinical and Medical Informatics in the Electronic Medical Record Era": Pre-Course_Questionnaire.pdf

### Quiz Instructions

This is the pre-course questionnaire, please fill this out so we can get to know you prior to you joining us in the course.

---

**Question 1** **0 pts**

What is your name (Firstname,Lastname): ______ , ______

**Question 2** **0 pts**

With what gender do you identify?

- Male
- Female
- Other

**Question 3** **0 pts**

What is your age?

______

**Question 4** **0 pts**

What is your current degree program: [degree]

#### Question 5

**0 pts**

Do you have a particular interest in a clinical specialty (due to research, or future or current practice)?

- Anesthesiology
- Dermatology
- Emergency Medicine
- Family Medicine
- Internal Medicine
- Med-Peds (Internal Medicine / Pediatrics)
- Neurosurgery
- Neurology
- Obstetrics & Gynecology
- Ophthalmology
- Orthopedic Surgery
- Otolaryngology (Ears, Nose, and Throat)
- Pathology
- Pediatrics
- Physical Medicine & Rehabilitation
- Plastic Surgery

- Psychiatry
- Radiation Oncology
- Radiology
- Surgery
- Urology

#### Question 6

**0 pts**

Have you previously participated in any research projects with a university or hospital environment?

- True
- False

#### Question 7

**0 pts**

Why did you select this elective? (You may select more than one)

- Previously described by former students as a bird course
- Offers a connection to the Regenstrief Institute
- Interest in data analytics
- Interest in big data
- Interest in medical administration

#### Question 8

0 pts

Please describe why you decided to pursue this elective

HTML Editor

12pt Paragraph

0 words

#### Question 9

0 pts

Do you plan on an academic/research career?

- True
- False

#### Question 10

0 pts

Do you plan on using informatics in your future practice?

True

False

#### Question 11 0 pts

What experience can we provide that would mean you find this elective worthwhile?

HTML Editor

12pt Paragraph

0 words

#### Question 12 0 pts

Define medical informatics

HTML Editor

12pt Paragraph

0 words

#### Question 13

**0 pts**

Define biomedical informatics.

HTML Editor

12pt Paragraph

0 words

**Question 14** **0 pts**

What problems do people in the field of medical informatics try to solve?

HTML Editor

12pt Paragraph

0 words

**Question 15** **0 pts**

How has the US government influenced the field of medical informatics?

HTML Editor

12pt Paragraph

0 words

#### Question 16

**0 pts**

List some of the barriers to Health Information Technology (HIT) adoption.

HTML Editor

12pt Paragraph

0 words

#### Question 17

0 pts

Describe the educational and career opportunities within medical informatics.

HTML Editor

12pt Paragraph

0 words

Saving... Submit Quiz
