## Supplemental File and Appendix for "Designing a Clerkship Curriculum for Medical Students in Clinical and Medical Informatics in the Electronic Medical Record Era": Syllabus.docx

### [Pre-Course Questionnaire](%24CANVAS_OBJECT_REFERENCE%24/quizzes/i234d6552f84faeeffd7973d22632d430)

This course consists of several modules, delivered in sequence, and in parallel. These modules consist of:

- Content intended to be consumed over a week long period, oriented towards self study

- Deliverables to be completed at the end of each week

Two one hour tutorial sessions will be made available with current fellows to support students working through problems with any of the modules they are working on. At the end of each week, students will be tasked to join the faculty and fellows in a discussion of the luminary paper series with a seminar discussion of the literature provided.

### Parallel

In parallel, there are three modules, run throughout the entire of the elective rotation:

**These modules all have deliverables during the course of the course, and must be completed by the end of the course**

1. The Luminary Paper Series

This module consists of readings from six luminary academics in the field of medical informatics, and their major contributions:

1. Larry Weed: The Problem-based Medical Record/Electronic Medical Record
2. Clement McDonald, Marc Overhage and Bill Tierney: The Electronic Medical Record/Network of Records (Health Information Exchange)
3. Edward Shortliffe: MYCIN
4. Octo Barnett: MUMPS
5. Naomi Sager and Carol Friedman: Natural Language Processing
6. Richard N Shiffman and Stephen M Downs: Clinical Guidelines and the Computer

As an additional resource, the Office of the Director of the National Library of Medicine hosts Dean Sittig and Joan Ash’s project documenting an oral history of [medical informatics pioneers](http://lhncbc.nlm.nih.gov/project/medical-informatics-pioneers).

2. Final Project

This module consists of several deliverables:

- Write design document
- Build a database
- Build a decision support module for their primary specialty
- Program the link between these two as a deliverable

3. Programming (with Python) and Databases

This module consists of an overview of object-oriented programming with the [Python](https://www.python.org/) programming language. For the programming component, exercises and content consists of working through the book Practical Programming (Second Edition): An Introduction to Computer Science Using Python 3, by *Gries, Campbell and Montojo*, and providing the instructors with the completed exercises. For the database component, the student is expected to complete Jennifer Widom’s course on databases on Stanford’s edX instance: [Lagunita](https://lagunita.stanford.edu/courses/DB/2014/SelfPaced/about), and upload the completion certificates to the instructors.

### Sequence

In sequence, students are expected to complete at a minimum of four week long modules, beginning with HIPAA training and ending with a choice between the three Regenstrief research foci modules:

**The student is expected to complete these modules in sequence, beginning with HIPAA Training**

1. HIPAA Training (Day 1)

Regenstrief requires students to go through additional HIPAA training than that provided by IU, and students will be required to complete both prior to continuing in the course. Additional readings are provided for a deep dive into some elements of HIPAA and related law and its impact on clinical or research practice. This module should not take a week to complete, it must be completed immediately, as you cannot access the first week’s module (Fundamentals of Medical Informatics) without completing this module.

2. Fundamentals of Medical Informatics

This module walks the student through a selected overview of the familiar fundamentals of medicine, and their application towards medical informatics. In addition, this module describes a brief history of medical and clinical informatics.

3. Decision Science

This module introduces the student to the fundamentals of decision science, as well as information theory, two core domains for medical informatics. It then describes how these two domains apply to the clinic, in terms of clinical decision support as well as clinical reasoning.

#### Regenstrief Research Foci Modules

These modules all represent major research topics of investigators at the Regenstrief Institute, and specific expertise that the institute has that outline particular elements of the domain of medical informatics.

**The student is expected to complete the terminology module during the course of the course**

4. Terminologies and Standards

This module provides an overview to standards in medicine, focusing on information-based standards and Logical Observation Identifiers Names and Codes (LOINC), a standard developed and maintained by the Regenstrief Institute. It specifically describes the following standards and services:

```
- The Unified Medical Language System (UMLS): Metathesaurus et al
    - The UMLS Terminology Services
- Systematized Nomenclature of Medicine - Clinical Terms (SNOMED-CT)
- International Classification of Diseases v9/10 (ICD-9/ICD-10)
- RxNorm
- Health Level 7 Messaging v2.8 and Fast Healthcare Interoperability Resources (FHIR)
- Logical Observation Identifiers Names and Codes (LOINC)
```

5. A Survey of Regenstrief’s Current Research

This module provides an overview of research currently being undertaken within the Center for Biomedical Informatics. Specifically, these fall under the realm of:

- Pubic Health Informatics
- The Teaching Electronic Medical Record
- Global Health Informatics and the Open Medical Record System
- Hospital and Clinical Operations
- Pediatric Informatics

### Attendance

If you are currently in the city, and do not have an excused absence, you are expected to attend the tutorial sessions, as well as luminary paper series lectures. Should you be away, the following information is available for you to dial in remotely. Please let us know of your absence by email.

From a web browser (we recommend Chrome), go to <https://bridge.iu.edu>, enter 236766777 and your name, then click Connect.

From Skype for Business, make a video or audio call to

From an IU videoconferencing room system, dial 236766777

From a non-IU videoconferencing room system, dial

From a telephone as an audio-only participant, dial (812)856-7060 or (317)278-8080, then 236766777

More connection information is available at <https://kb.iu.edu/d/ause>. Need help? Contact the IU Video Help Desk at (812) 856-2020,.

### Grading

Grading in Canvas primarily counts towards your participation and engagement in the course material.

The learning management system also serves as a means to track your reading activity and a preliminary measurement of depth of inquiry for each week’s in-person content.

As with other electives, this course is graded at the discretion of the course director (Dr. Finnell).

This elective is primarily intended to build and assess your problem solving skills through the integration of medical informatics knowledge and content into your prospective clinical practice and care delivery.

Your grade will reflect your understanding of the material in this context and demonstration of the translation of said knowledge into examples demonstrating fealty with evidence-based medicine and models of disease built upon scientific and clinical knowledge.

In order to obtain a passing grade in this elective, you must complete the bare minimum of content (following assignments):

- Integrating Terminologies for Diagnosis/Disease Classification
- Modeling a Decision Problem
- Design Document Deliverable
- Final Report Deliverable

Your engagement with the materials will determine your obtention of honours or high honours. That said, it will be favourably looked upon if you complete additional assignments, and the depth at which you do so will be reflected in your grade. Furthermore, for those who indicate a predilection towards research, these assignments offer and content offer a way to engage with the institute in more depth, and gain base-level skills expected of researchers in clinical informatics.

The United Nations Educational, Scientific and Cultural Organization (UNESCO) Institute of Statistics has released the 2011 International Standard Classification of Education. In this system, the American Medicinae Doctor degree is equivalent a ISCED Level 7 degree: Second or further degree programme at Master’s or equivalent level (following successful completion of a Bachelor’s or equivalent programme). Specifically:

United States — First-professional degree programmes. Completion of these programmes signifies both completion of the academic requirements to begin practicing in a given profession and a level of professional skill beyond that normally required from a Bachelor’s degree. These programmes typically last three years and require at least two years at ISCED level 6 prior to entrance (although most require a four-year Bachelor’s degree). These first professional degrees are the normal route to qualify as a professional in the fields of dentistry, medicine, optometry, pharmacy, veterinary medicine, law and theological professions.

To this end, to guide you in your readings, we would refer you to advice provided in the Chronicle for Higher Education <http://chronicle.com/article/A-Letter-to-Past/236870> by Rachel Herrmann PhD, a lecturer in early modern American history on the Faculty of Humanities at the University of Southampton. This advice applies globally to the readings and general requirements of graduate level courses, and although the MD is not a ISCED Level 8 degree (that of the doctorate); we expect a level of effort commensurate with the notion that you are effectively in graduate school rather than undergraduate.
